# Multimodal Machine Learning Reveals the Genomic and Proteomic Architecture of Heart Failure with Preserved Ejection Fraction

**DOI:** 10.64898/2026.02.07.26345811

**Authors:** Jack W. O’Sullivan, Taedong Yun, Ruoyi Cai, David Amar, Tim Assimes, Akshay Chaudhari, Dan S. Kim, Eldrin Lewis, Francois Haddad, Farhad Hormozdiari, J Weston Hughes, Gabriel Mannis, Michael Salerno, Mark Pepin, James Pirruccello, Jared Wallace, Howard Yang, Manuel Rivas, Andrew Carroll, Cory Y. McLean, Euan A. Ashley

**Affiliations:** Division of Cardiology, Department of Medicine, Stanford University, CA, USA; Department of Biomedical Data Science, Stanford University, CA, USA; Google Research, Cambridge, MA, USA; School of Computer Science and AI, Tel Aviv University, Israel; VA Palo Alto Healthcare System, Palo Alto, CA, USA; Department of Radiology, Stanford University, CA, USA; Division of Cardiology, Department of Medicine, University of Washington, Seattle, WA, USA; Division of Hematology and Oncology, Department of Medicine, Stanford University, CA, USA; Division of Cardiology, Department of Medicine, University of California, San Francisco, CA, USA; Division of Cardiology, Department of Medicine, Columbia University, NY, USA; Google Research, Mountain View, CA, USA

**Author notes:** Co-first authors (Jack W. O’Sullivan, Taedong Yun). Co-last authors, order determined randomly (Cory Y. McLean, Euan A. Ashley).

## Abstract

Heart failure with preserved ejection fraction (HFpEF) affects over 30 million people and lacks disease-modifying therapies. Although genomic-led drug discovery increases success by more than 2.6-fold, HFpEF genomic discovery remains constrained by imprecise phenotyping in biobanks, with only two loci identified to date. Biobanks lack HFpEF diagnostic codes and echocardiograms, yet HFpEF diagnosis exists along a continuum and is inherently probabilistic, presenting an opportunity for multimodal prediction.

Here we introduce TRI-modal Assessment and Discovery of HFpEF (TRIAD-HFpEF), a machine learning framework integrating electrocardiograms, cardiac magnetic resonance imaging, and biomarkers to assign HFpEF probabilities. Deployed in UK Biobank, these probabilities validate with respect to mortality, hospitalizations, and structural and functional HFpEF features.

Genome-wide and proteomic analyses reveal over 90 novel loci, a 45-fold expansion, and distinguish causal proteins from non-causal biomarkers of disease progression, prioritizing 11 therapeutic targets and 7 non-causal biomarkers. We identify FLT3 as one of the 11 therapeutic targets, consistent with the reported 7-fold increased heart failure risk from FLT3 inhibitors in leukemia. We validate this finding by demonstrating significant worsening of diastolic function following FLT3 inhibitor treatment in an independent clinical cohort. Conversely, MPO emerged as one of the 7 non-causal biomarkers, aligning with three recent negative MPO inhibitor trials. TRIAD-HFpEF demonstrates that machine learning-derived phenotypes can unlock genetic discovery in complex syndromes, identifying actionable targets while deprioritizing associations reflecting disease consequences rather than causes.

## Introduction

Heart failure with preserved ejection fraction (HFpEF) affects over 30 million people, costs $30 billion annually in the US alone, and carries a 30% mortality rate within one year of hospitalization.^1–4^ Current therapies reduce heart failure hospitalizations but have limited impact on mortality.^3–7^ As genomic-led drug discovery has demonstrated a 2.6-fold improvement in successful drug development,^8^ there have been “urgent” calls to understand the genetic architecture of HFpEF.^9^ Despite this, our current understanding of the genetic determinants of HFpEF is poor: the two prior dedicated genome-wide association studies (GWAS) have discovered only two, marginally significant loci (*FTO* and *IGFBP7*).^9,10^

HFpEF diagnosis requires evidence of diastolic dysfunction (assessed exclusively on echocardiograms) and clinical signs of volume overload via diuretic use and elevated natriuretic peptides (NT-proBNP/BNP).^11,12^ Even with these data readily available, HFpEF diagnosis in clinical practice is often probabilistic, with the disease existing across a broad clinical spectrum and probabilistic tools frequently used clinically.^13,14^ Prior efforts to elucidate the genomic and molecular mechanisms of HFpEF have been hindered by non-specific disease coding in genetic biobanks.^15–18^ Global biobanks are unable to assign binary HFpEF disease codes due to the absence of echocardiograms, NT-proBNP or BNP data, and incomplete medication records. As a result, there are no HFpEF codes in UK Biobank (UKB), and all heart failure cases are coded non-specifically as “congestive heart failure,” “left ventricular failure,” or “heart failure, unspecified.”

Despite the inability to assign binary HFpEF disease codes in UKB, the rich availability of cardiac data in UKB and the probabilistic nature of HFpEF diagnosis presents a potential opportunity to predict HFpEF from electrocardiograms (ECG), cardiac magnetic resonance imaging (CMR) and biomarkers. Recent advances in machine learning have led to this approach being successfully applied to other diseases with similarly continuous and often probabilistic diagnoses.^19^ Such approaches derive disease probabilities from multimodal clinical data, effectively transforming noisy or incomplete diagnostic signals into quantitative disease phenotypes. Such “digital twin” approaches have been employed, providing genetically informative traits that enhance power for downstream discovery.^20,21^

Here we present TRI-modal Assessment and Discovery of HFpEF (TRIAD-HFpEF), a machine-learning (ML) framework to predict HFpEF from ECGs, CMRs, and biomarkers. TRIAD-HFpEF models were trained in a large clinical dataset and then deployed to UKB to assign HFpEF probabilities to participants. TRIAD-HFpEF-assigned predicted probabilities were then used as inputs for genomic and proteomic analyses, ultimately discovering over 90 novel loci, prioritizing 11 HFpEF therapeutic targets, and 7 non-causal disease-progression biomarkers. We open-source all model code, weights, and molecular findings to further advance the field. Our findings provide novel insights into the molecular mechanisms underlying HFpEF, and present an array of potential targets for further therapeutic investigation.

## Methods

The study consisted of four main phases: 1. The construction of precise HFpEF phenotypes for machine learning models using real-world clinical data (at Stanford), 2. The training of ML models to predict HFpEF from ECG (TRIAD-ECG), CMR (TRIAD-CMR), and biomarkers (TRIAD-LAB), 3. Deployment of these models in UKB to assign HFpEF probabilities for all participants and validate predictions, 4. Genomic and multi-omic analyses in UKB using predicted HFpEF probabilities (Figure 1). The study was approved by Stanford’s Institutional Review Board (IRB: 67663, 41045) and under UKB project ID 22282.

**Figure 1.**
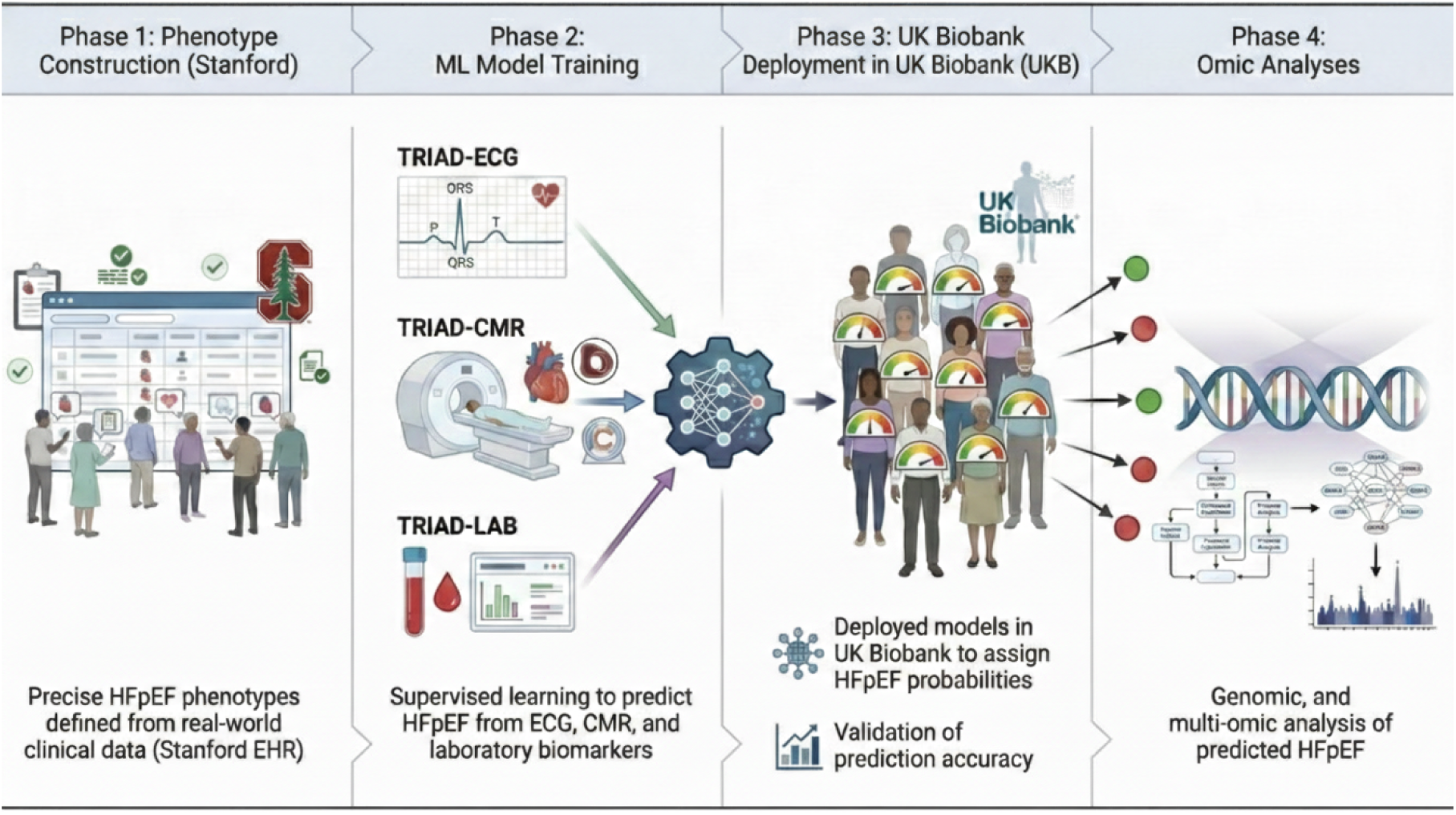
Study design. **A)** Training of ML models in a clinical cohort (Stanford). 3 individual models were trained, validated and tested. **B)** Deployment of these models into UKB, where participants were assigned a HFpEF probability. **C)** Multi-omic analyses performed in UKB using HFpEF probabilities as inputs.

### Stanford dataset

Real-world clinical data used for ML model training were obtained from Stanford health care, a large tertiary health care system hospital in Northern California. All data were routinely collected for clinical care and then deidentified for research use. Institutional Review Board approval was obtained (41045 and 67663). We identified all Stanford patients that had undergone an echocardiogram and a) an ECG and b) a CMR. We had access to raw ECG and CMR images, as well as physician text reports of these images. Similarly, we had access to demographic and clinical data in tabular form (e.g. medication use, laboratory values).

### Construction of precise HFpEF phenotype

We defined HFpEF cases within the Stanford data as either a) patients with a HFpEF electronic health record (EHR) code or, b) in the absence of a HFpEF EHR code, patients who had an echocardiogram with a preserved LVEF (>50%), abnormal diastolic function, NT-proBNP or BNP elevation, and diuretic use (in line with international guidelines^12,22^). To determine diastolic function, we constructed regular expressions to extract key measures from echocardiogram data (left ventricular ejection fraction, diastolic measures: lateral e’, medial e’, mitral E/A ratio, E/e’ ratio, tricuspid regurgitation (TR) velocity, and left atrium (LA)-volume index).^23^ We followed the American Society of Echocardiogram (ASE) guidelines^23^ and defined evidence of diastolic dysfunction as two or more of: E/e’ ratio>14, lateral e’<10 cm/s or medial e’<7 cm/s, TR velocity>2.8 m/s, and LA-volume index>34 ml/m^2^).^23^ To create more granular phenotypes, we also calculated each patient’s H2FPEF score: a well-validated score that uses age, body mass index (BMI), E/e’ ratio, pulmonary artery systolic pressure, and atrial fibrillation history to give H2FPEF probability.^13^ Patients with a history of cardiac transplant, congenital heart disease, and pulmonary arterial hypertension were excluded. The phenotyping code is available open-source at: https://github.com/jackosullivanoxford/HFpEF_phenotyping/code_to_phenotype_HFpEF.

### Training ML models: TRIAD-HFpEF

We designed and trained three separate machine learning models to predict HFpEF: TRIAD-ECG to predict HFpEF from ECGs, TRIAD-CMR to predict HFpEF from CMRs, and TRIAD-LAB to predict HFpEF from clinical biomarkers.

#### TRIAD-ECG

From Stanford, we identified all patients that had undergone an ECG and an echocardiogram. For all these patients we used the phenotyping approach (‘*Construction of Precise HFpEF Phenotype’)* to classify patients as HFpEF case (1) or control (0) at Stanford. For all of these patients, the ECG that was closest to their echocardiogram date was selected and converted to matrices with shapes representing leads (12) and data points (5000: representing a sampling rate of 500 over 10 seconds). Matrices were normalized across leads and baseline and notch filters were applied. These matrices were used as input to our deep learning model, which consisted of multiple convolutional blocks with progressively increasing filter counts similar to our prior work,^24^ enhanced by systematic alternation between channel attention and spatial attention mechanisms integrated throughout the convolutional layers (see Supplementary for full model architecture description, Supplementary Figure 1, https://github.com/jackosullivanoxford/TRIAD_ECG_HFPEF for code and https://huggingface.co/jackosullivan/TRIAD_ECG_HFPEF for weights). The Stanford ECGs were split into train, test and validation, with class imbalance hyper-parameter set to 0.20. We used the Adam optimizer with an initial learning rate of 1×10⁻⁵, weight decay of 1×10⁻³, and batch size of 32 for up to 1000 epochs. We employed a ReduceLROnPlateau scheduler with a patience of 15 epochs and minimum learning rate of 1×10⁻⁷ (reduction factor of 1×10⁻²). Our model was trained on a single Nvidia Xp GPU using python 3.9 and PyTorch 1.11. The probabilities produced from this model were then incorporated with 117 numeric ECG measurements reflecting intervals (such as PR interval), and amplitudes (e.g. T wave amplitude) using XGBoost.

#### TRIAD-CMR

We identified all patients at Stanford who had undergone a CMR from 2005 to 2023 and extracted imaging DICOMs representing three standard views: 4-chamber view (horizontal long axis), 3-chamber view, and 2-chamber view (vertical long-axis) (Supplementary Figure 2). Among all patients for whom all three views of CMR are available, we selected patients who had also undergone echocardiography. The target phenotype we used for TRIAD-CMR was two-fold: we assigned patients as having HFpEF (or not) using the above phenotyping (which used regular expressions to extract LVEF from echocardiogram, diuretic use and NT-proBNP/BNP) and also a continuous phenotype from the H2FPEF score.

We identified only the cines (videos) of the three views that are static in location but vary in time (4 chamber/Horizontal long axis, 2 chamber/vertical long axis, and 3 chamber). Each cine is assumed to represent a single cardiac cycle as it is standard for CMR, starting from the end of the diastole. For consistency, all cines were preprocessed to have exactly 8 images per cine and the cines with fewer than 8 images were discarded.

To train our deep learning TRIAD-CMR model, we augmented the training and validation dataset by applying up to ±10° rotations (−10, 5, 0, 5, 10) and minor changes in image brightness (−2%, 0%, 2%, 4%, 6%), resulting in 121825 (= 4873 × 25) training cases and 15225 (= 609 × 25) validation cases. We adapted the widely-used ResNet V2 model for three cines (24 images) by applying the model to each of the 24 images with an additional logistic regression layer using all 24 outputs. The shared ResNet V2 weights and the weights of the final logistic regression layer were jointly trained using the continuous HFpEF probability labels we developed, the binary cross entropy loss (with the “soft” probability labels), and AdamW optimizer for 128 epochs, starting from the ImageNet checkpoint available in Keras and TensorFlow2. The model with the minimal validation set loss was selected as our final TRIAD-CMR model.

#### TRIAD-LAB

All patients that had an echocardiogram at Stanford were identified. For these patients, we then identified laboratory results all routinely collected at Stanford (complete blood count, complete metabolic panel, and lipid panel, Supplementary Table 1), and relevant clinical and demographic variables (such as age, sex, BMI etc). Prior work suggests that HFpEF can be predicted with good accuracy from a parsimonious selection of clinical factors, namely: age, BMI, and history of atrial fibrillation.^25^ We first confirmed a similar accuracy in predicting HFpEF from these three clinical factors in our Stanford dataset (test AUC 0.81), and then in three open-source datasets with very similar results to prior work (Supplementary Figure 3).^25^ Similar to the MILTON approach,^20^ we then extended this model with the above 39 covariates (Supplementary Table 1). All training inputs were standardized to mean 0 and unit variance. We used XGBoost with hyperparameter tuning to predict HFpEF from these covariates. We split data into train, valid, and test cohorts. All code and model weights are available open-source https://github.com/jackosullivanoxford/TRIAD_LAB_HFpEF for code and https://huggingface.co/jackosullivan/TRIAD_LAB_HFPEF for weights.

### Deployment of TRIAD-HFpEF to UK Biobank

All covariates and data sources used to train TRIAD-HFpEF models (ECG, CMR, and LAB) are available at both Stanford and in UKB. We followed the same pre-processing steps to clean the CMR and ECG data described above (e.g. baseline and notch filters for ECGs). We used the model weights from our trained TRIAD-HFpEF models and deployed these trained models to UKB (TRIAD-ECG used UKB ECGs as inputs, TRIAD-CMR used UKB CMRs as inputs and TRIAD-LAB used UKB biomarkers as inputs). We deployed each of these models individually, which generated a predicted probability of HFPEF for participants in UKB.

#### Validation of HFpEF predicted probabilities

To examine the validity of our HFpEF predicted probabilities in UKB we conducted the following analyses: 1. We determined the correlation between our HFpEF predicted probabilities and the numeric values available for CMR (e.g. left ventricle mass, left atrial volume), 2. We determined the accuracy of our TRIAD models in predicting HFpEF probabilities in patients with a left ventricular ejection fraction (LVEF) >50% on CMR and *not* predicting HFpEF in patients with an LVEF <50%, 3. We conducted survival analyses using our HFpEF predicted probabilities as inputs to predict a) CV mortality, b) Non-specific heart failure (HF) ICD code, and c) HF hospitalization. All models were adjusted for age and sex, and open-source code is available here: https://github.com/jackosullivanoxford/HFpEF_phenotyping

### Genome-wide association studies and genetic heritability

The above three models (TRIAD-LAB, TRIAD-ECG, and TRIAD-CMR) assigned a probability of HFpEF to UKB participants, respectively (TRIAD-LAB N=502,134, TRIAD-ECG N=81,607, and TRIAD-CMR N=84,433). To maintain the unique biological signals identified by each separate model, we conducted three separate genome-wide association studies (GWAS) for each of the probabilities assigned by these three models, treating them as three continuous traits for GWAS. Standard GWAS quality controls were applied. For each of the GWAS summary statistics produced from our three analyses, we calculated heritabilities and genetic correlation between all three traits using linkage disequilibrium score regression.^26^ Further, we obtained GWAS summary statistics from the prior HFpEF GWAS in the Million Veterans Program (MVP),^9^ which identified one genome-wide significant locus (FTO), and conducted genetic correlation with our results. We used PLINK and BOLT-LMM for genome-wide association studies. We then estimated the genetic heritability of TRIAD-ECG, TRIAD-CMR, and TRIAD-LAB individually and the genetic correlation between each pair of GWAS results and the MVP HFpEF GWAS.

### Proteomic analyses

We conducted the following proteomic analyses in UKB: Proteome-wide association study, bidirectional Mendelian randomization using multiple methods (inverse-variance weighted (IVW) and MR Egger), and Protein colocalization.

### Proteome-wide association study

We used linear models to assess the strength of association between HFpEF predicted probabilities and protein expression. We defined strength of association by regression coefficient in all statistically significant associations. For the proteomic analyses, the code is available open source at: https://github.com/jackosullivanoxford/HFpEF_phenotyping/Proteome_wide_analysis.

### Protein quantitative trait locus (pQTL) Mendelian randomization and colocalization analysis

We conducted bidirectional Mendelian randomization (MR) analyses. For the “forward” direction (protein → HFpEF), we used 1Mb cis-protein quantitative trait loci (cis-pQTLs) as instrumental variables, with protein levels as the exposure using summary statistics from UKB-Pharma Proteomics Project^27^ and HFpEF predicted probability as the outcome using our TRIAD HFpEF GWAS (3 analysis respectively for TRIAD-LAB, TRIAD-ECG, and TRIAD-CMR). For the “reverse” direction (HFpEF → protein), we used TRIAD HFpEF GWAS variants as instruments, with HFpEF liability as the exposure and protein levels as the outcome. The inverse-variance weighted (IVW) method^28^ was employed as the primary analysis and MR Egger^29^ as sensitivity analysis. All reported associations were subsequently corrected for multiple testing using the False Discovery Rate (FDR) method. Lastly, we performed colocalization to identify shared pathways between significant variants identified from GWAS and pQTL results using COLOC^30^ (Version 5.2.3) within R (Version 4.5.1). Finally, we prioritized the most likely gene-protein pathways as proteins that were significant in the proteome-analysis, forward pQTL MR, and colocalization. Code for all analyses is available open source (See Supplementary: *All open source code)*.

### Clinical Validation of FLT3

Our integrative analyses identified 11 potential HFpEF therapeutic targets, including FLT3, for which higher expression appears protective against HFpEF. This finding generates a testable prediction: pharmacological FLT3 inhibition should worsen diastolic function, the hallmark of HFpEF, while sparing systolic function. FLT3 inhibitors are routinely used in leukemia, presenting a natural experiment. Prior studies have reported a 7-fold increased heart failure risk with these agents, though the mechanism and heart failure subtype remain uncharacterized. We therefore identified all patients at Stanford treated with FLT3 inhibitors who had pre- and post-treatment echocardiograms, extracting diastolic parameters and left ventricular ejection fraction. Comparing pre- and post-treatment function while adjusting for time between echocardiograms, age, and anthracycline exposure allowed us to test whether FLT3 inhibition specifically impairs diastolic function consistent with HFpEF pathophysiology.

## Results

### Construction of Precise HFpEF Phenotype

To create phenotypes for our TRIAD-ECG and TRIAD-LAB model training, we identified 115,149 patients at Stanford that had undergone an ECG and an echocardiogram. Of these 115,149 patients, 11,276 (9.8%) patients were classified as HFpEF and 103,873 (90.2%) as controls (in line with expected hospital prevalence^31^) (Figure 2A). Our regex-based model extracted all diastolic parameters (median values reported in Supplementary Table 2) and we identified the ECG taken at the closest time point to the echocardiogram date (median days between ECG and echocardiogram: 0).

**Figure 2.**
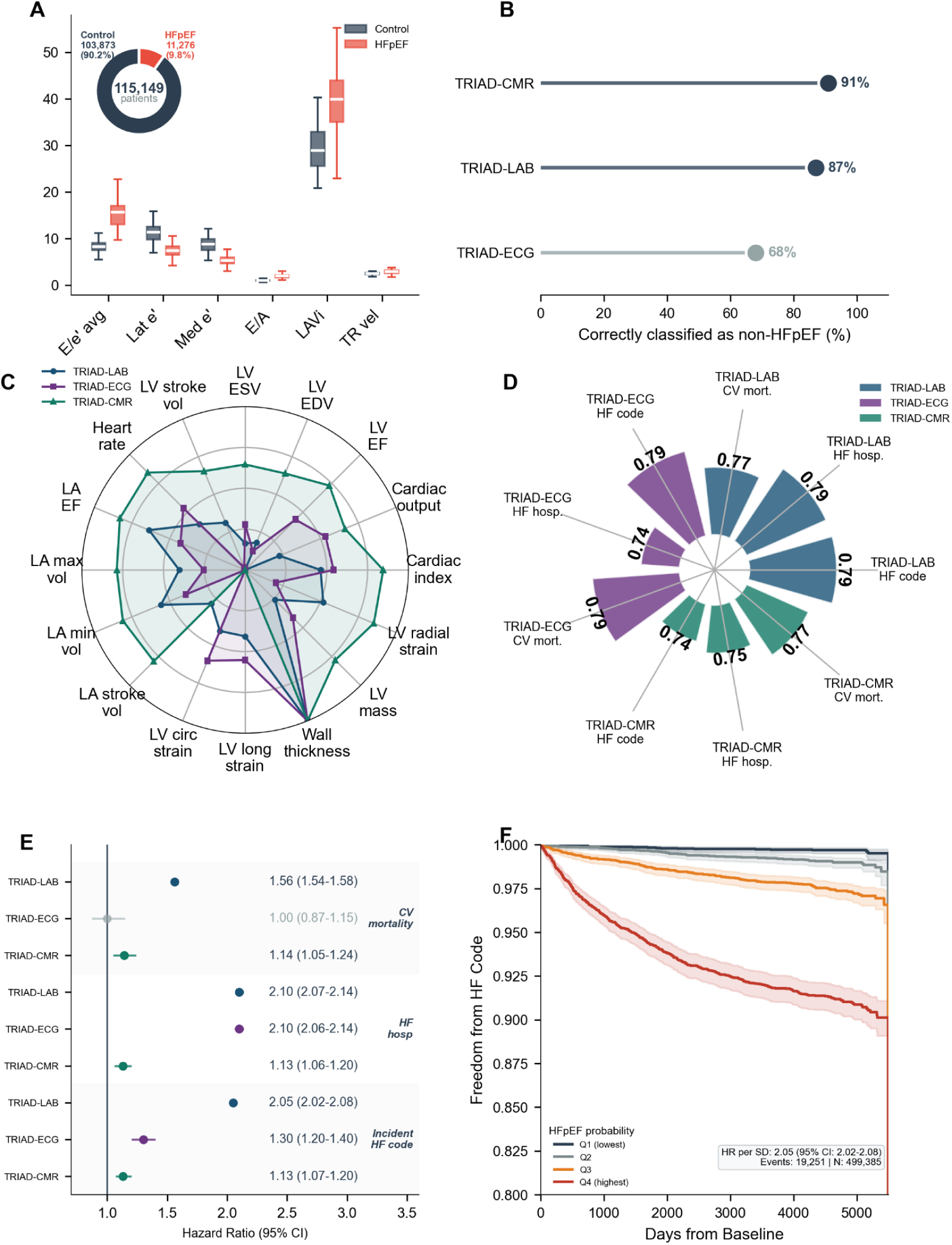
Case/control distribution at Stanford and validation of TRIAD-HFpEF models in UKB. **(A)** Stanford training cohort (n=115,149) composition and diastolic parameters comparing HFpEF cases (red) versus controls (blue). E/e’, ratio of early mitral inflow velocity to mitral annular velocity; Lat e’, lateral mitral annular velocity; Med e’, medial mitral annular velocity; E/A, ratio of early to late mitral inflow velocity; LAVi, left atrial volume indexed; TR vel, tricuspid regurgitation velocity. **(B)** Specificity for HFpEF: percentage of UK Biobank participants with left ventricular ejection fraction (LVEF) <50% correctly classified as non-HFpEF. **(C)** Log-normalized associations between predicted HFpEF probabilities and cardiac magnetic resonance features. LV, left ventricular; LA, left atrial; EF, ejection fraction; EDV, end-diastolic volume; ESV, end-systolic volume. **(D)** C-statistics for predicting incident heart failure (HF) code, HF hospitalization and cardiovascular (CV) mortality. **(E)** Hazard ratios (95% CI) per s.d. increase in predicted HFpEF probability. Grey indicates non-significant associations. **(F)** Kaplan–Meier curves for incident HF by TRIAD-LAB probability quartiles (n=499,385; 19,251 events; median follow-up 13.9 years).

To create phenotypes for our TRIAD-CMR, we identified 6,092 patients who had all three CMR views of the cine (3 × 8 = 24 images per patient) and a label for HFpEF, whom we divided into three groups, training (80%; N=4,873), validation (10%; N=609), and test (10%; N=610). We augmented this data as described in the methods and thus ultimately had 121,825 training cases and 15,225 validation cases.

### Training and Accuracy of TRIAD-HFpEF Models

We successfully trained three models to predict HFpEF from ECG (TRIAD-ECG), CMRs (TRIAD-CMR) and biomarkers (TRIAD-LAB). The respective area under the receiving order curve (AUC) for the held out test set for the three models was: TRIAD-ECG: 0.81, TRIAD-CMR: 0.74, and TRIAD-LAB: 0.84.

### Deployment of TRIAD-HFpEF models and validation of phenotypes

We deployed our three TRIAD-HFpEF models into UKB and assigned a predicted probability of HFpEF from each model. There were 502,134 UKB participants with available biomarker data for the deployment of our TRIAD-LAB model, 81,607 with ECG data and 84,439 with CMR data. There are no HFpEF specific disease codes or echocardiograms for ground truth labeling in UKB. To ensure the accuracy of our HFpEF predicted probabilities in UKB, we compared our predicted probabilities with structural and functional features present in HFpEF. We validated predicted HFpEF probabilities via their association with expected structural HFpEF changes (Figure 2C). Left atrial (LA) ejection fraction showed the strongest association (p < 1×10^-200^ for TRIAD-LAB; p = 2.5×10⁻⁶⁵ for TRIAD-ECG, p = 5.7×10⁻^3^⁵ for TRIAD-CMR), with lower LA ejection fraction corresponding to higher HFpEF risk, consistent with known impairments in LA function in HFpEF. LA maximum volume (p = 4.5×10⁻¹⁰⁴ for TRIAD-LAB; p = 1.3×10⁻⁴² for TRIAD-ECG, p = 2.3×10⁻^39^ for TRIAD-CMR) was also positively associated with HFpEF probability, reflecting larger atrial volumes associated with HFpEF in keeping with known physiology. Similarly, HFpEF probabilities were significantly associated with lower LV end-diastolic volume (p = 7.4×10⁻¹⁶ for TRIAD-LAB; p = 2.8×10⁻¹⁹ for TRIAD-ECG, p = 3.1×10^-4^ for TRIAD-CMR) and end-systolic volume (p = 2.9×10⁻⁵ for TRIAD-LAB; p = 4.4×10⁻³⁹ for TRIAD-ECG, p = 1.5×10^-2^ for TRIAD-CMR), higher myocardial mass (p = 1.1×10⁻¹² for TRIAD-LAB; R² = 0.0039, p = 2.1×10⁻³¹ for TRIAD-ECG, p = 5.4×10^-29^ for TRIAD-CMR), and increased myocardial thickness (p = 1.6×10⁻²⁰ for TRIAD-LAB; p = 1.1×10⁻³⁶ for TRIAD-ECG, p = 1.3×10^-158^ for TRIAD-CMR). These significant associations are consistent with HFpEF, despite the absence of imaging data as training inputs for both TRIAD-LAB and TRIAD-ECG. We also validated the accuracy of our models to correctly identify LVEF <50% as not HFpEF cases. TRIAD-LAB classified 87% of these participants correctly as *not* HFpEF cases (Figure 2B). TRIAD-ECG correctly classified 68% of participants as not HFpEF and TRIAD-CMR 91%.

### Survival analysis

We further validated our predicted HFpEF probabilities in UKB via their association with a) Non-specific HF code, b) HF hospitalization, and c) CV mortality.

Predicted HFpEF probabilities from TRIAD-ECG, TRIAD-CMR, and TRIAD-LAB all significantly predicted HF hospitalization, and non-specific HF code. TRIAD-LAB derived HFpEF probabilities (N=502,128) predicted incident HF code: HR: 2.05 (95%CI: 2.02-2.08, p<0.00005, C-statistic: 0.79), HF hospitalization HR: 2.10, 95%CI: 2.07-2.14, p<0.00005, C-statistic: 0.79), and CV mortality HR: 1.56, 95%CI: 1.54-1.58, C-statistic: 0.77). For the 81,536 participants with ECG data in UKB, we found that TRIAD-ECG HFpEF probabilities significantly predicted incident HF ICD code (HR: 1.3, 95%CI: 1.2-1.4, p=2e-9, C-statistic: 0.79), and HF hospitalization (HR: 2.10, 95%CI:2.06-2.14, p<0.000005, C-statistic: 0.74), but was for CV mortality the HR was not significant (HR: 1.0, 95%CI: 0.9-1.2, C-statistic: 0.79). For the 84,433 participants with CMR data, TRIAD-CMR significantly predicted incident HF code (HR: 1.13 95%CI:1.07-1.20 p<0.00003, C-statistic: 0.74), HF hospitalization (HR: 1.13 95%CI:1.06-1.20, p<0.00008, C-statistic: 0.75), and CV mortality: (HR: 1.14 95%CI:1.05-1.24, p=0.02, C-statistic: 0.77).

### Genome-wide association studies

Across the three modality-specific GWAS (TRIAD-LAB, TRIAD-ECG, and TRIAD-CMR), we identified 101 independent genome-wide significant loci (P < 5×10⁻⁸), revealing both shared and distinct components of HFpEF genetic architecture (Table 1, full GWAS summary statistics available open source, see supplementary). Convergent signals across modalities implicated metabolic regulation (FTO, PRKAG2), cardiac development and electrophysiology (SOX5, CASZ1), and extracellular matrix and inflammatory pathways (THBS3, SAA2–SAA4). Notably, the FTO locus on chromosome 16q12.2 emerged as genome-wide significant in all three analyses, highlighting a central metabolic axis in HFpEF pathogenesis and providing some validation of our methodological approach. These cross-modality findings suggest that ML-derived phenotypes from laboratory, ECG, and imaging data capture complementary dimensions of HFpEF biology: metabolic, structural, and electrical.

**Table 1.** Top annotated hits from genome-wide association studies.

| Chr:Pos Ref/Alt | rsID | Nearest Gene(s) | $-\log_{10}(p)$ |
| --- | --- | --- | --- |
| <u>TRIAD LAB</u> |  |  |  |
| 9: 113,925,534 T/C | rs7023923 | AL162414.1 | 40.587 |
| 2: 182,315,885 G/T | rs7573465 | ITGA4 | 36.895 |
| 16: 53,806,453 A/G | rs56094641 | FTO | 36.312 |
| 7: 151,415,536 C/T | rs10254101 | PRKAG2 | 29.588 |
| 1: 155,178,782 A/T | rs760077 | THBS3, MTX1 | 24.793 |
| <u>TRIAD ECG</u> |  |  |  |
| 12: 24,599,578 A/G | rs10842350 | SOX5 | 9.089 |
| 6: 117,479,490 A/G | rs4120420 | RNA5SP214 | 8.107 |
| 2: 179,711,376 T/C | rs2220127 | AC092640.1, CCDC141 | 8.044 |
| 16: 53,805,207 C/A | rs11075985 | FTO | 7.863 |
| 18: 57,846,077 C/T | rs663640 | RNU4-17P | 7.8 |
| <u>TRIAD CMR</u> |  |  |  |
| 16:53,800,954 T/C | rs1421085 | FTO | 15.11 |
| 11:18,263,463 GAATA/G | - | SAA2-SAA4,SAA2 | 9.04 |
| 15:74,783,298 C/G | rs182786760 | UBL7 | 7.3 |

The TRIAD-LAB GWAS identified 93 independent lead SNPs reaching genome-wide significance (P < 5×10⁻⁸); many near genes with established roles in metabolic and inflammatory regulation (e.g., FTO, PRKAG2, ITGA4). Notably, FTO and PRKAG2) emerged as top hits, implicating metabolic dysregulation as a central HFpEF mechanism (Table 1, Figure 3).

**Figure 3:**
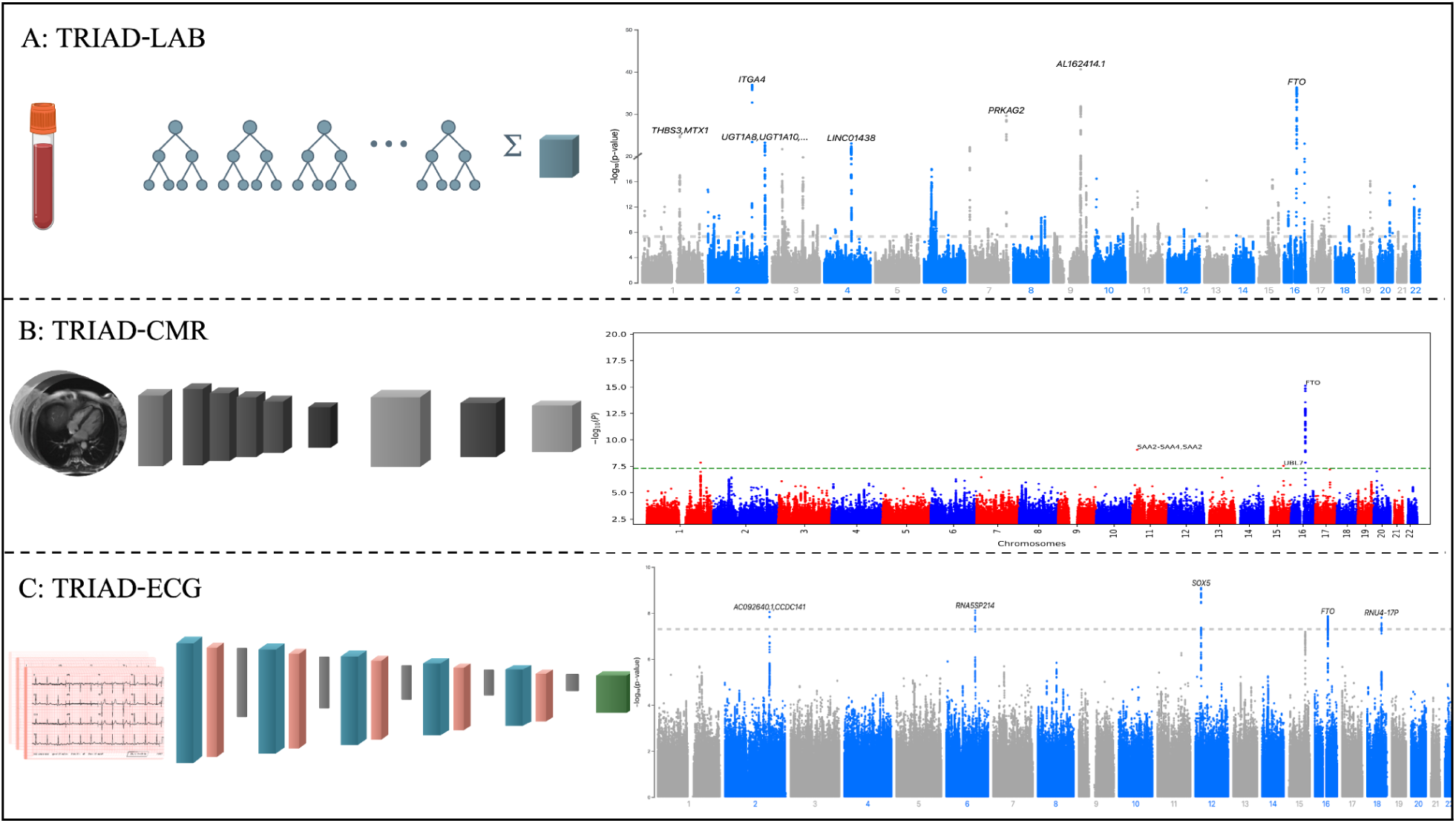
GWAS Manhattan plots. **(A)** GWAS from TRIAD-LAB. **(B)** GWAS from TRIAD-CMR. **(C)** GWAS from TRIAD-ECG.

The TRIAD-ECG GWAS identified five independent loci surpassing genome-wide significance (P < 5×10⁻⁸), implicating genes central to cardiac development and metabolic regulation (Table 1). The top association mapped to SOX5 (12q13.2), a transcription factor critical for cardiac chamber morphogenesis and conduction system formation. Additional significant signals included CCDC141, linked to sinoatrial node function, and FTO, replicating the metabolic association observed in TRIAD-LAB. Collectively, these findings suggest that ECG-derived phenotyping captures both developmental and metabolic axes of HFpEF susceptibility

The TRIAD-CMR GWAS yielded three independent loci exceeding genome-wide significance (P < 5×10⁻⁸), dominated by a robust signal at FTO (16q12.2; Table 1). This locus showed the strongest effect across all three TRIAD modalities, reinforcing a shared metabolic underpinning of HFpEF pathogenesis. Additional associations near SAA2–SAA4 and UBL7 implicated acute-phase inflammatory signaling and ubiquitin-mediated protein turnover, respectively. Together, these findings suggest that CMR-derived phenotypes capture metabolic and inflammatory components of HFpEF, complementing the developmental and structural pathways highlighted by TRIAD-LAB and TRIAD-ECG.

Together, these three GWAS reveal a convergent genetic framework for HFpEF centered on metabolic, inflammatory, and developmental pathways. The recurrence of FTO across all modalities underscores the metabolic dimension of disease risk, while modality-specific signals in SOX5 and SAA2–SAA4 point to distinct cardiac developmental and inflammatory processes.

### Genetic heritability

Using the predicted HFpEF probabilities in the UKB, we estimated the heritability of HFpEF to be around 9% (TRIAD-LAB (9.1%), TRIAD-CMR (9.4%), TRIAD-ECG (8.6%), Supplementary Figure 4) and genomic inflation factors up to 1.36 (TRIAD-LAB: 1.36, TRIAD-CMR: 1.05, TRIAD-ECG: 1.09). There was minimal population stratification bias observed (LDSC intercept of 1.04 for TRIAD-LAB, 1.003 for TRIAD-CMR, and for 1.023 TRIAD-ECG).

The genetic correlations between all three TRIAD GWAS results was significant. The genetic correlation between TRIAD-LAB and TRIAD-CMR was 0.498 (SE = 0.073, p = 1.01 × 10^-11^), for TRIAD-LAB and TRIAD-ECG, it was 0.602 (SE = 0.069, p = 1.58 × 10^-18^) and for TRIAD-ECG and TRIAD-CMR 0.729 (SE = 0.130, p = 1.79 × 10^-8^).

The genetic correlation between the TRIAD-LAB and TRIAD-CMR GWAS results and the MVP GWAS results was also significant. The genetic correlation between TRIAD-LAB and MVP was 0.261 (SE = 0.058, p = 6.0 × 10^-6^), and 0.215 (SE = 0.067, p = 0.0013) for TRIAD-CMR and MVP. The genetic correlation for TRIAD-ECG and MVP was 0.173 (SE = 0.095), but was not significant (p = 0.069). Notably, the MVP GWAS had a small sample size (total N ∼20k) and did not perform detailed precision phenotyping.

### Proteome-wide association study

To further decipher the molecular basis of HFpEF, we extended our analysis from genomics to proteomics. There were 2,940 proteins available for analysis in UKB. Our TRIAD-LAB proteome-wide association study revealed significant associations between HFpEF and 1969 plasma proteins (adjusted p<0.05), (Figure 4A, Supplementary table 3). Leptin (LEP) showed the strongest positive association (β=1.63, FDR<10⁻³⁰⁰), consistent with the established obesity-HFpEF link, followed by natriuretic peptide precursor B (NTPROBNP: β=1.30, FDR<10⁻³⁰⁰) and B-type natriuretic peptide (NPPB: β=1.19, FDR=3.8×10⁻¹⁷⁰). Other highly significant associations included oxytocin (OXT: β=1.20, FDR=1.8×10⁻¹⁴⁴), fatty acid-binding protein 4 (FABP4: β=1.03, FDR<10⁻³⁰⁰), renin (REN: β=1.02, FDR<10⁻³⁰⁰), and glucagon (GCG: β=1.01, FDR=6.3×10⁻¹⁴¹). Additional notable associations included gastrin (GAST: β=0.97, FDR=2.6×10⁻¹²²) and fatty acid-binding protein 1 (FABP1: β=0.82, FDR=2.0×10⁻¹⁴⁸). Insulin-like growth factor binding protein 1 (IGFBP1: β=-0.74, FDR=1.1×10⁻⁶⁷) demonstrated a strong negative association, suggesting potential protective mechanisms related to insulin sensitivity. These protein associations remained significant after adjusting for age, sex, genetic ancestry, and batch effects (Supplementary Table 3). Results were similar for TRIAD-ECG (Supplementary Table 4, Supplementary Figure 6) and TRIAD-CMR (Supplementary Table 5, Supplementary Figure 7)

**Figure 4:**
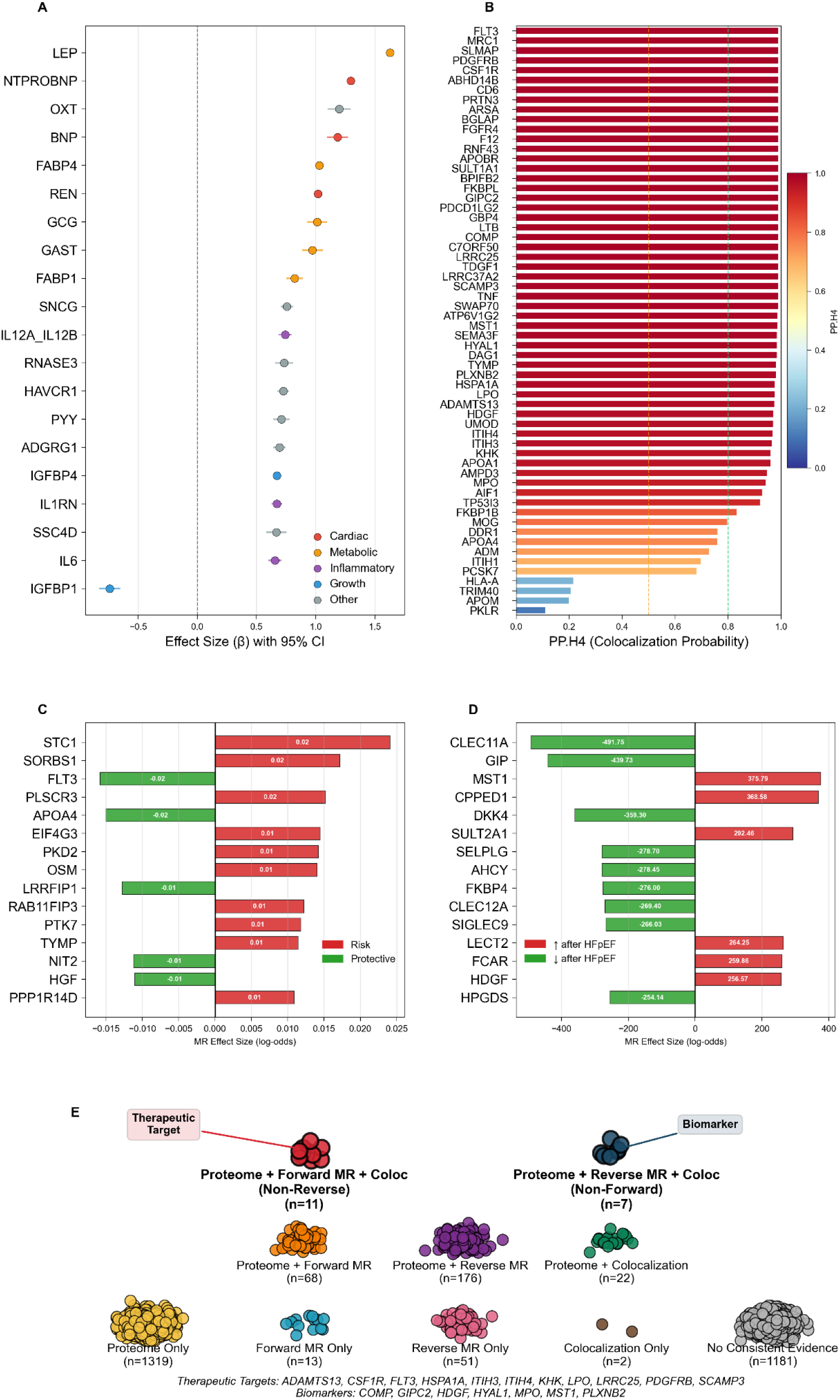
Gene-Protein results for TRIAD-LAB. **(A)**: Results from proteome-wide association study: 20 proteins with expression levels most strongly associated with HFpEF predicted probability. **(B)**: Colocalization results: Proteins with highest Posterior Probability of Hypothesis 4 (PP.H4), listed in highest to lowest. **(C)**: Forward Mendelian Randomization (MR) results: green indicates higher protein expression levels are protective of HFpEF, red risk enhancing. **(D):** Reverse Mendelian Randomization results: green indicates protein expression decreases after the development of HFpEF, red indicates protein expression increases after development of HFpEF. **(E):** Identifying potential therapeutic gene-protein pathways and non-casual gene-protein pathways that likely represent biomarkers. Therapeutic targets are significant across protein-wide (‘Proteome’), Forward Mendelian Randomization (‘Forward MR’), and Colocalization (‘Coloc’) and not Reverse Mendlian Randomization. Biomarkers are significant across protein-wide (‘Proteome’), Reverse Mendelian Randomization (‘Forward MR’), and Colocalization (‘Coloc’) and not Forward Mendlian Randomization.

### Colocalization analysis

To further identify gene-protein pathways dysregulated in HFpEF, we performed colocalization analysis to identify shared genomic and proteomic signals at HFpEF risk loci. For our TRIAD-LAB colocalization analysis we identified 50 proteins with significant pQTL-HFpEF colocalization evidence (PP.H4 > 0.80; the posterior probability that two traits share a same causal variant), suggesting that genetic variants affecting circulating protein levels share causal mechanisms with HFpEF susceptibility (Figure 4B, Supplementary Table 5, Supplementary Figure 8). The proteins with the highest PP.H4 probabilities were: FLT3, MRC, SLMAP, PDGFRB, and CSF1R.

For our TRIAD-ECG and TRIAD-CMR analysis given the smaller number of GWAS hits, there were 10 proteins (Supplementary Table 7) and 0 proteins respectively that had pQTLs that overlapped with GWAS hits - none of which had a PP.H4 probability >0.80.

### Protein Quantitative Trait Loci Mendelian Randomization

We performed bidirectional pQTL Mendelian randomization (MR) to identify proteins potentially causal in HFpEF pathogenesis (forward MR) and proteins with expression altered as a consequence of HFpEF (reverse MR). First, we excluded proteins that were not significantly associated with any SNPs: 233 proteins had no significant SNPs. From the 2,707 remaining proteins we then extracted only cis-pQTLs using a ±1 Mb window around gene regions; 657 proteins had no cis hits leaving 2050 proteins for primary pQTL MR analysis.

Forward direction Mendelian randomization (protein -> HFpEF) TRIAD-LAB analysis identified 1,654 proteins with significant causal effects on HFpEF risk (adjusted FDR < 0.05) using the Inverse-Variance Weighted (IVW) (Supplementary Table 3). We prioritized proteins with concordant causal estimates across both IVW and MR-Egger methods to minimize pleiotropy and identified 229 proteins that were significant across both IVW and MR Egger (and had a non-significant intercept term) (Supplementary Table 8, Supplementary Figure 9). Of these 229 proteins, 170 were significant exclusively with forward MR (protein -> HFpEF) and not significant for reverse MR (HFpEF -> protein, Supplementary table 9, Figure 4C). For TRIAD-ECG, there were 1,492 proteins significant using IVW and 240 using both IVW and MR Egger (and had a non-significant intercept term). Of these 240 significant proteins, 179 were significant only in forward MR and not in reverse MR (Supplementary table 10, Supplementary Figure 11). For TRIAD-CMR, there were 685 proteins significant using IVW only and 84 using both IVW and MR Egger (and had a non-significant intercept term, Supplementary table 11, Supplementary Figure 13).

For reverse direction Mendelian randomization (HFpEF -> protein) TRIAD-LAB analysis we identified 259 proteins that were significant using both IVW and MR Egger (and had a non-significant intercept term (Supplementary table 12, Supplementary Figure 10). Of these 259 proteins, 199 were only significant in reverse direction analysis (Figure 4D). For TRIAD-ECG and TRIAD-CMR analyses, we identified 296 and 178 proteins respectively that were significant using both IVW and MR Egger (and had a non-significant intercept term, Supplementary Table 13 and Supplementary Table 14). Of these 296 proteins significant from TRIAD-ECG, 235 were significant only in the reverse analysis (Supplementary Figure 12). Similarly, of these 178 proteins from TRIAD-CMR, 129 were significant only in the reverse analysis (Supplementary Figure 14).

### Systematic prioritization of gene-protein pathways

To identify potential gene-protein pathways that have the highest likelihood of being dysregulated in the pathogenesis of HFpEF we identified proteins that met all of the following criteria for strong evidence: significant and concordant effect sizes across proteome-wide analysis, colocalization (PP.H4 > 0.80), and Egger and IVW pQTL forward MR, as well as not significant in the reverse MR pQTL analysis. We identified 11 proteins for TRIAD-LAB (Figure 4E, Supplementary table 15): most notably FLT3, CSF1R and PDGFRβ. For TRIAD-ECG and TRIAD-CMR we did not identify any proteins with colocalization PP.H4 > 0.80 hence we did not identify proteins meeting all four criteria, but we did identify 59 and 11 proteins significant and concordant across proteome-wide and forward pQTL for TRIAD-ECG (Supplementary Figures 17, 18) and TRIAD-CMR respectively (Supplementary Figure 19, 20).

Similarly, the reverse pQTL MR enabled discovery of causal proteins dysregulated after the onset of HFpEF could represent novel HFpEF biomarkers. We identified 7 proteins that meet all criteria for strong evidence (same as above except the opposite for forward and reverse) (Figure 4, Supplementary table 16). Most notably *MPO*, *MST1*, and *HDGF*.

### Clinical Validation of FLT3

The above systematic prioritization of gene-protein pathways identified 11 potential therapeutic targets including FLT3. Our results suggest genetically predicted higher FLT3 expression conferred protection from HFpEF. This generated a testable clinical hypothesis: FLT3 inhibition leads to diastolic dysfunction and HFpEF. Clinically, FLT3 inhibitors are widely used in acute myeloid leukemia presenting a natural experiment: does diastolic function worsen after FLT3 inhibitor use. To test this hypothesis, we identified 104 patients at Stanford who had been treated with FLT3 inhibitors and had pre and post treatment echocardiograms. We extracted LVEF, and 6 diastolic markers (lateral e’, medial e’, Mitral E/A ratio, left atrial volume index (LAVi), E/e’ average, Tricuspid velocity). After adjusting for time (between echocardiograms), age and concurrent anthracycline use, we noted a significant decrease in lateral e’, medial e’, and significant increase in E/e’ consistent with worsening diastolic dysfunction, with no significant change in LVEF, and no significant change in TR velocity, LAVi and Mitral E/A ratio (Extended Table 1, and Figure 5). Lateral and medial e’ deterioration are the earliest markers of diastolic dysfunction and HFpEF,^23^ whereas LAVI and TR velocity represent structural, later stage changes of diastolic dysfunction, and mitral E/A patterns are non-linear and should not be interpreted in isolation.^23^

**Figure 5:**
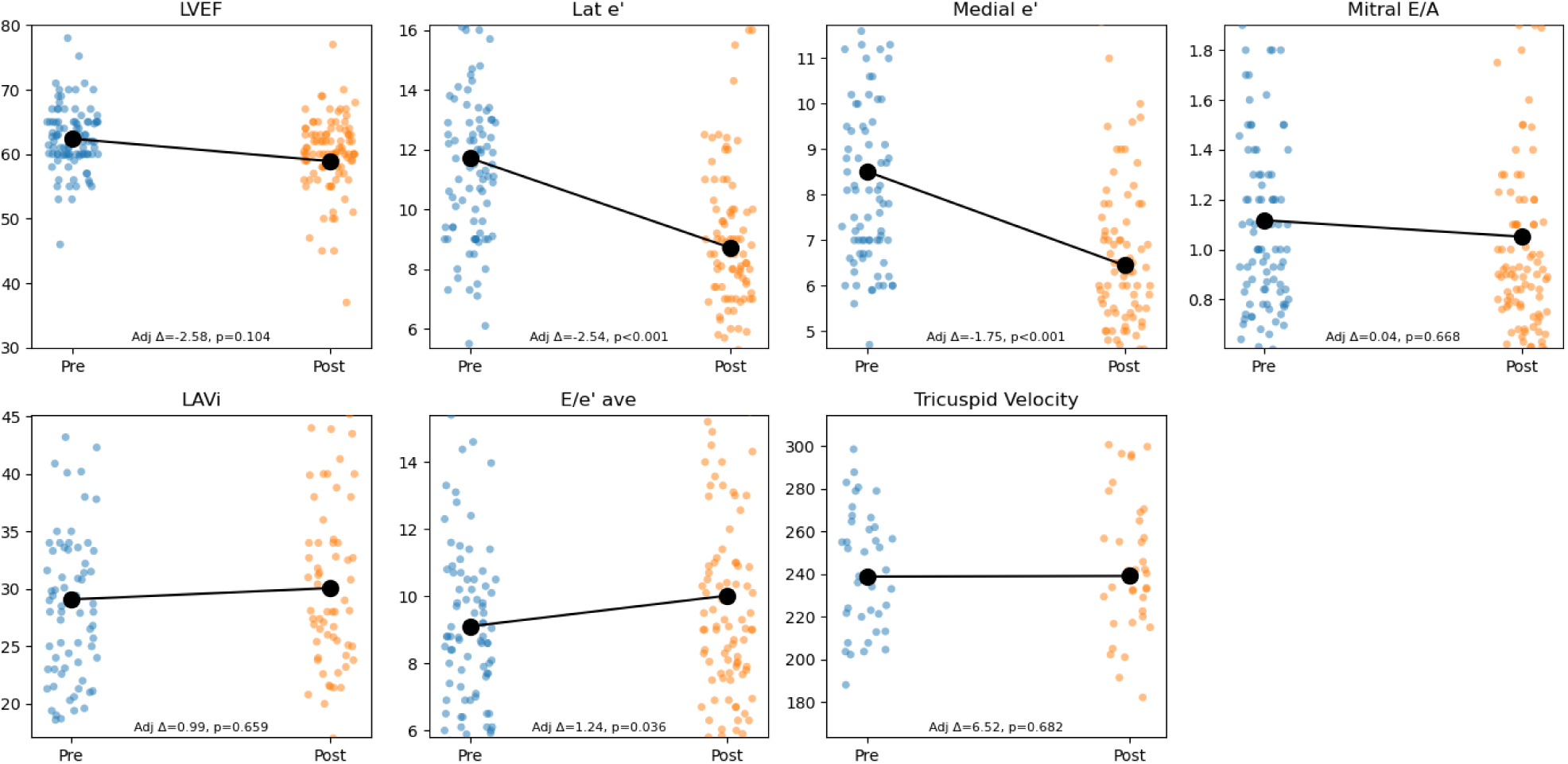
LVEF and diastolic markers pre and post FLT3 inhibitor treatment. Black dots repesent mean.

## Discussion

We present TRIAD-HFpEF, three ML models leveraging multimodal ECG, CMR, and biomarker data to predict HFpEF and elucidate its genomic and proteomic architecture. Our three significant contributions are: a) ML models to predict HFpEF from three common but distinct clinical data sources (ECGs, CMRs, and biomarkers), b) almost 100 novel HFpEF loci, a 45-fold increase in genomic discovery for HFpEF, and c) a refined list of casual therapeutic targets for HFpEF, with validation of one target in an independent cohort, and conversely a list of gene-protein pathways dysregulated as a consequence of HFpEF; more likely to represent biomarkers.

Our ML models were trained using a large-scale dataset from a US academic medical center, achieving high predictive accuracy. The models were then independently deployed in UK Biobank to generate continuous HFpEF phenotypes. These phenotypes capture the full spectrum of disease severity while incorporating the uncertainty inherent in probabilistic predictions. The predicted probabilities were validated via strong associations with established HFpEF features, including structural markers (e.g., increased min and max left atrial volumes), functional characteristics (e.g., preserved left ventricular ejection fraction), and outcomes (e.g., heart failure hospitalization). Using these validated predictions, we conducted genome-wide association studies, identifying 99 novel HFpEF loci (101 total). We then nominated and prioritized causal gene-protein pathways dysregulated in HFpEF, highlighting potential therapeutic targets. We identify 11 promising gene-protein pathways that have therapeutic potential with supporting validation and 7 gene-protein pathways that have biomarker potential. For a highly morbid condition lacking disease-modifying therapies, TRIAD-HFpEF represents the first comprehensive multi-omic discovery enabled by a multimodal machine learning framework.

Given the absence of truly disease modifying medications for HFpEF, we employed a genomic and multi-omic approach to identify gene-protein molecular pathways. Results revealed novel, untested molecular pathways, and pathways with existing therapeutic targets currently being investigated for other cardiac diseases. We prioritized potential therapeutic targets via tiered levels of evidence - from being significant only in GWAS, to the most compelling targets significant and concordant across GWAS, proteome-wide association, colocalization, and pQTL MR. Across the TRIAD-LAB GWAS, the *PRKAG2* pathway, represents a molecular pathway with a currently investigated therapy that could be reproposed for HFpEF. Monogenic variants in *PRKAG2* cause excessive glycogen storage in the heart via AMP-activated protein kinase (AMPK) activity. The clinical phenotype shares similarities with HFpEF via left ventricular hypertrophy, diastolic dysfunction and preserved LVEF. There are small interfering RNA (siRNA) inhibitors of *PRKAG2* that have been shown to improve diastolic function and left atrial remodeling, which suggests that perhaps *PRKAG2* siRNAis could be efficacious in HFpEF.^32^

Beyond genome-wide signals, our integrative proteogenomic analyses identified 11 proteins strongly associated with the pathogenesis of HFpEF, supported by proteome-wide analyses, protein colocalization, and forward MR. These dysregulated proteins represent potential novel therapeutic targets for HFpEF. The observed protective effect of FLT3^33–36^ is supported by preclinical, real-world studies, randomized controlled trials (RCTs), and our own clinical validation data. FLT3 activation preserves myocardial homeostasis in multiple preclinical models: FLT3 signalling regulates cardiac side-population progenitors and capillary maintenance, and exogenous FLT3 activation mitigates stress-induced cardiomyocyte hypertrophy and fibrosis through SIRT1/p53-dependent preservation of mitochondrial dynamics (e.g., OPA1) and autophagy.^33–36^ These mechanistic data provide a plausible route by which higher FLT3 activity would protect against the microvascular rarefaction, energetic dysfunction and maladaptive remodelling that typify HFpEF. FLT3 inhibitors are used in the treatment of acute myeloid leukemia and have consistent cardiotoxic side effects. A 2023 real-world analysis showed patients on FLT3 inhibitors have an almost 7-fold increased risk of heart failure cardiac toxicity.^37^ Similarly, FLT3 inhibitor RCTs have shown increased risk of heart failure incidence, death and heart failure symptoms.^38^ The ADMIRAL RCT compared Gilteritinib, a FLT3 inhibitor, with conventional chemotherapy (without an FLT3 inhibitor) and found the Gilteritinib group had significantly higher rates of peripheral edema, dyspnea, and congestive cardiac failure.^38^ Lasty, our validation cohort within Stanford’s leukemia patients confirmed significant worsening of diastolic function after FLT3 inhibitors without any effect on systolic function. Collectively, our integrative analyses establish FLT3 as a cardioprotective mediator in HFpEF, with independent preclinical, randomized, and real-world studies providing orthogonal evidence of worsening diastolic function and HFpEF with FLT3 inhibition. While the therapeutic potential of FLT3 activation in HFpEF remains unclear, the multitiered evidence suggesting FLT3 inhibiton causes diastolic dysfunction and HFpEF validates our integrative approach to identify HFpEF therapeutic targets.

The systematic priorization of gene-protein pathways also distinguished causal proteins (therapeutic targets) from those altered as disease consequences (biomarkers). Among the most promising biomarker candidates, MPO and HDGF emerged with robust supporting evidence from both our integrative analyses and prior independent work. MPO is an emerging HFpEF biomarker and therapeutic target that has been investigated extensively. Our results suggest that MPO protein levels are increased after the development of HFpEF, representing a biomarker not a therapeutic target. Prior RCTs support this conclusion: the 2024 ENDEAVOR HFpEF trial examined mitiperstat, a *MPO* inhibitor, and showed no benefit across primary nor secondary outcomes.^39^ Similarly, the 2024 SATELLITE HFpEF trial showed no difference in NT-proBNP levels, 6 minute walk distance, symptoms, or HFpEF echocardiogram findings with an MPO inhibitor^40^ and the 2025 Popovic randomized double blinded HFpEF trial showed no reduction in exertional hemodynamic congestion with MPO inhibition.^41^ Despite the lack of clinical trial therapeutic benefit, prior observational work confirms biomarker potential with significantly elevated MPO levels in HFpEF patients compared to controls and strong correlations with established disease markers including NT-proBNP and diastolic dysfunction (E/e’ ratio).^42,43^ Taken together, this integrative, multitiered approach identified MPO as a downstream biomarker of HFpEF, a conclusion strengthened by concordant findings from independent randomized and real-world investigations.

Collectively, this novel, integrative, multitiered proteogenomic approach identified candidate therapeutic targets validated across orthogonal lines of evidence (including FLT3), while also identifying pathways that appear to be downstream consequences of HFpEF, representing potential biomarkers that should not be pursued for therapeutic development.

Our study has limitations. There was no ground truth for our HFpEF predictions in the UKB. This is a limitation inherent in the approach, and similar digital twin methodologies. We attempted to mitigate this in two ways: First, we used an entirely independent training cohort, which had precision phenotype definitions. Prior work that employed a similar ‘digital twin’ approach^20,46^ constructed models and deployed them in the same cohort. For example, in MILTON, all training, deployment of models to identify digital twins and follow up GWAS occurred in one cohort (UKB).^20^ Second, we validated our HFpEF predicted probabilities in three ways: (1) Association with structural and functional HFpEF features: we found that our HFpEF predicted probabilities were strongly associated with known HFpEF features, such as left atrial volumes (min and max), left ventricular wall thickness, and myocardial mass, (2) Classification by left ventricular ejection fraction (LVEF): tabular data on LVEF is available on ∼85,000 participants. This presented an opportunity to identify participants with an LVEF <50% and thus definitively do not have HFpEF. Our models correctly classified 68-91% of participants as not HFpEF cases (LVEF <50%), and (3) Association with non-specific heart failure codes, heart failure hospitalization, and cardiovascular mortality. All HFpEF predictions from TRIAD-LAB, TRIAD-CMR, and TRIAD-ECG were strongly associated with non-specific heart failure codes and hospitalization, Further, some of the non-novel genomic and proteomic results further validate our predictions. Across all three GWAS, we re-identified the only consistently significant HFpEF locus (FTO), and in our protein-wide analyses we showed that natriuretic peptides NT-proBNP and BNP (biomarkers integral to HFpEF diagnosis) and leptin had the strongest protein-wide associations with HFpEF predicted probability, closely followed by Renin. A further limitation is potential inaccuracy in our model-derived HFpEF probabilities. HFpEF diagnosis exists along a biological and clinical spectrum;^13,14^ higher predicted HFpEF probability values (e.g., 0.9) from our models indicate greater confidence in the presence of underlying pathobiology and reflect more advanced or definitive phenotypic expression. Further, while individual predicted probabilities may not be precisely calibrated (for example, 0.75 vs. 0.76), the relative ranking of participants is preserved. This rank ordering is critical for downstream genomic analyses, where association strength depends on relative, not absolute, phenotypic differences. In fact, the use of a continuous HFpEF phenotype may be a strength of our approach. Continuous phenotypes inherently captures uncertainty and are less erroneous than clinical binary codes.^47^ A 2020 study examined the accuracy of heart failure codes to identify HFpEF and found that codes were accurate only 50% of the time, and ‘should not be used alone’.^47^

To conclude, TRIAD-HFpEF is the first multimodal machine learning framework to predict HFpEF and comprehensively map its genomic and proteomic landscape. TRIAD-HFpEF achieved high predictive accuracy and enabled continuous HFpEF phenotyping that was validated through strong associations with established disease features and outcomes. This framework facilitated the largest HFpEF genome-wide association study to date, expanding known genetic loci from 2 to 101 and revealing novel pathophysiological pathways. The integrative analyses identified 11 promising therapeutic targets (notably *FLT3*) and 7 biomarker candidates (notably *MPO*).

## Acknowledgments

This research has been conducted using the UK Biobank Resource under Application Numbers 22282 and 65275.

## Data Availability Statement

The UKB data used in this study are available via the usual research application procedures to UK Biobank. The Stanford data was accessed under an IRB and an anonymized subset is being released open-source upon peer-review publication.

