## Supplementary information for "Multimodal Machine Learning Reveals the Genomic and Proteomic Architecture of Heart Failure with Preserved Ejection Fraction"

### Methods

*Training ML models: TRIAD-HFpEF*

TRIAD-ECG

The network architecture employed a dual-attention CNN design with multiple convolutional blocks and progressively increasing filter counts: 64, 128, 256, 512, and 1024 filters. Each convolutional layer used 1D kernels of varying sizes ((1, 15), (1, 11), or (1, 7)) with appropriate padding to maintain feature map dimensions. The key architectural innovation was the systematic alternation between channel attention and spatial attention mechanisms after each convolutional layer. Channel attention modules identified the most informative feature channels using global average pooling followed by a squeeze-and-excitation mechanism, while spatial attention modules highlighted important temporal regions using 7×7 convolutions with sigmoid activation. Max-pooling layers with kernel sizes of (1, 2) or (1, 4) provided temporal downsampling between blocks. The architecture concluded with an adaptive average pooling layer and fully connected layers (1024 units) with batch normalization, ReLU activation, and dropout regularization.

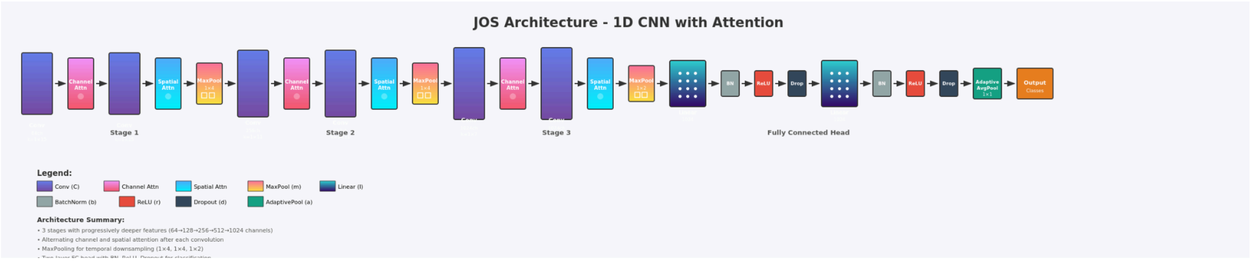


**Supplementary Figure 1: Full 'JOS' architecture of model for TRIAD-ECG prediction: Convolutional Neural Network with attention layers.**

TRIAD-CMR

Given the limitations in sample size for the CMR cohort we used the continuous H2FPEF phenotype, the median H2FPEF score was 14.2% (IQR: 0-46.6%).

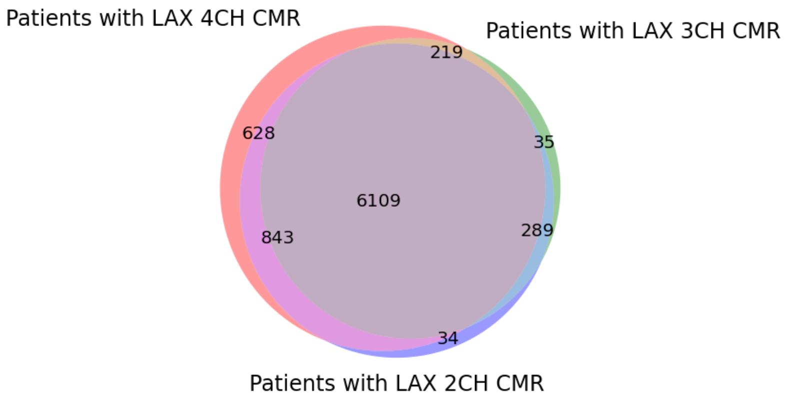


**Supplementary Figure 2 The number of patients with each type of CMR.**

TRIAD-LAB

*Validation of prior HFpEF-ABA score*

The below is the accuracy metrics of our provisional model validated on four open-source external datasets from Reddy et al.[25]

| Dataset | N | Cases | Case% | Accuracy | Precision | Recall | F1 | AUC |
|---|---|---|---|---|---|---|---|---|
| Derivation ABA score Final | 414 | 267 | 64.50% | 0.742 | 0.801 | 0.798 | 0.799 | 0.809 |
| Hospitalized HFpEF validation | 629 | 456 | 72.50% | 0.804 | 0.821 | 0.934 | 0.874 | 0.912 |
| International Validation ABA | 736 | 563 | 76.50% | 0.739 | 0.833 | 0.824 | 0.829 | 0.766 |
| Second validation cohort | 228 | 179 | 78.50% | 0.728 | 0.868 | 0.771 | 0.817 | 0.765 |

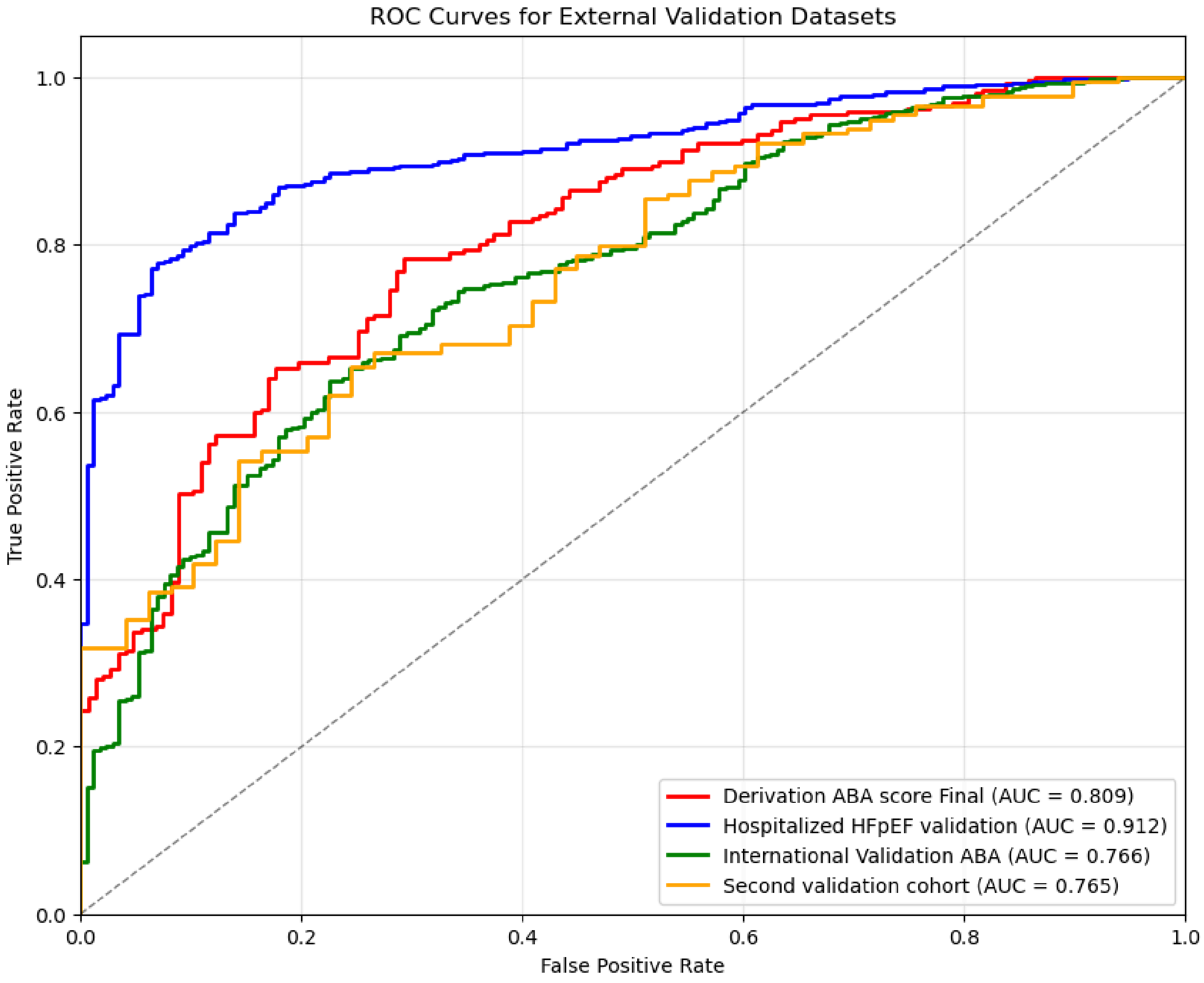


**Supplementary Figure 3:** Receiver Operator Curve (ROC) for simple model predicting HFpEF from Age, BMI, and history of atrial fibrillation.

*Training ML Models: TRIAD-HFpEF*

<u>TRIAD-ECG</u>

The tabular 117 ECG measurements used were:  III_pamp', 'III_pdur', 'III_qamp', 'III_qdur', 'III_ramp', 'III_rdur', 'III_samp', 'III_sdur', 'III_tamp', 'II_pamp', 'II_pdur', 'II_qamp', 'II_qdur', 'II_ramp', 'II_rdur', 'II_samp', 'II_sdur', 'II_tamp', 'I_pamp', 'I_pdur', 'I_qamp', 'I_qdur', 'I_ramp',, 'I_rdur', 'I_samp', 'I_sdur', 'I_tamp', 'V1_pamp', 'V1_pdur', 'V1_qamp',  'V1_qdur', 'V1_ramp', 'V1_rdur', 'V1_samp', 'V1_sdur',  'V1_tamp', 'V2_pamp', 'V2_pdur', 'V2_qamp', 'V2_qdur', 'V2_ramp', 'V2_rdur', 'V2_samp', 'V2_sdur', 'V2_tamp', 'V3_pamp', 'V3_pdur', 'V3_qamp', 'V3_qdur', 'V3_ramp', 'V3_rdur',  'V3_samp', 'V3_sdur', 'V3_tamp', 'V4_pamp', 'V4_pdur', 'V4_qamp', 'V4_qdur', 'V4_ramp', 'V4_rdur', 'V4_samp', 'V4_sdur',  'V4_tamp', 'V5_pamp', 'V5_pdur', 'V5_qamp', 'V5_qdur', 'V5_ramp', 'V5_rdur', 'V5_samp', 'V5_sdur', 'V5_tamp', 'V6_pamp', 'V6_pdur', 'V6_qamp', 'V6_qdur', 'V6_ramp', 'V6_rdur', 'V6_samp', 'V6_sdur', 'V6_tamp', 'aVF_pamp', 'aVF_pdur', 'aVF_qamp', 'aVF_qdur', 'aVF_ramp', 'aVF_rdur',

'aVF_samp', 'aVF_sdur',  'aVF_tamp', 'aVL_pamp', 'aVL_pdur', 'aVL_qamp', 'aVL_qdur', 'aVL_ramp', 'aVL_rdur', 'aVL_samp', 'aVL_sdur', 'aVL_tamp', 'aVR_pamp', 'aVR_pdur', 'aVR_qamp', 'aVR_qdur', 'aVR_ramp', 'aVR_rdur', 'aVR_samp', 'aVR_sdur', 'aVR_tamp', 'meanqrsdur', 'meanqtc', 'meanqtint', 'meanrrint', 'meanventrate', 'pfrontaxis', 'qrsfrontaxis', 'tfrontaxis', 'avg_pdur
Key: pamp = p-wave amplitude, pdur = p-wave duration, qamp = q-wave amplitude, qdur=q-wave duration, ramp= R-wave amplitude, rdur = R-wave duration, samp = S wave amplitude, sdur = S wave duration, tamp = T-wave amplitude, avg_pdur = average P duration
ECG leads: I, II, III, aVL, aVR, aVF, V1-V6.

### Results

*Genetic heritability*

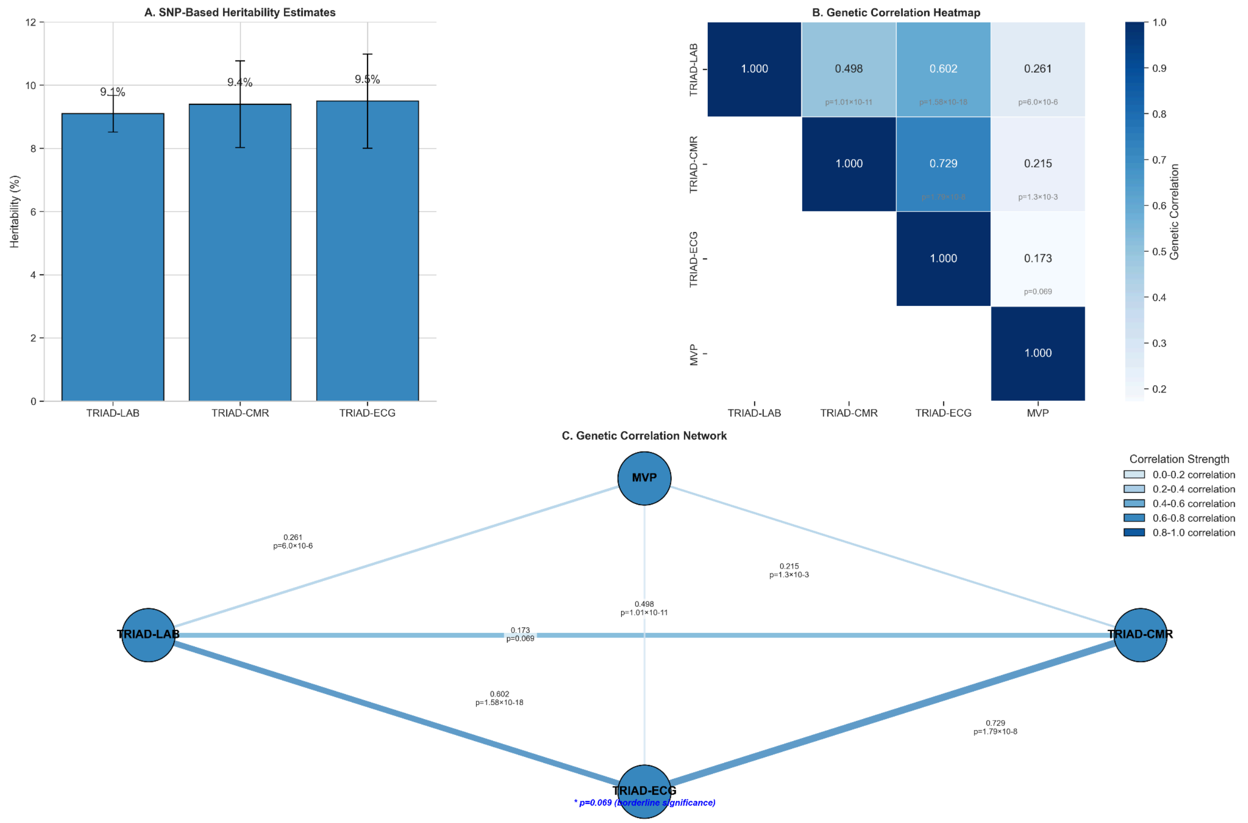


**Supplementary Figure 4** Genetic heritability estimates between 3 TRIAD-HFpEF analyses and MVP HFpEF GWAS.

*Proteome-wide analysis*

TRIAD-LAB

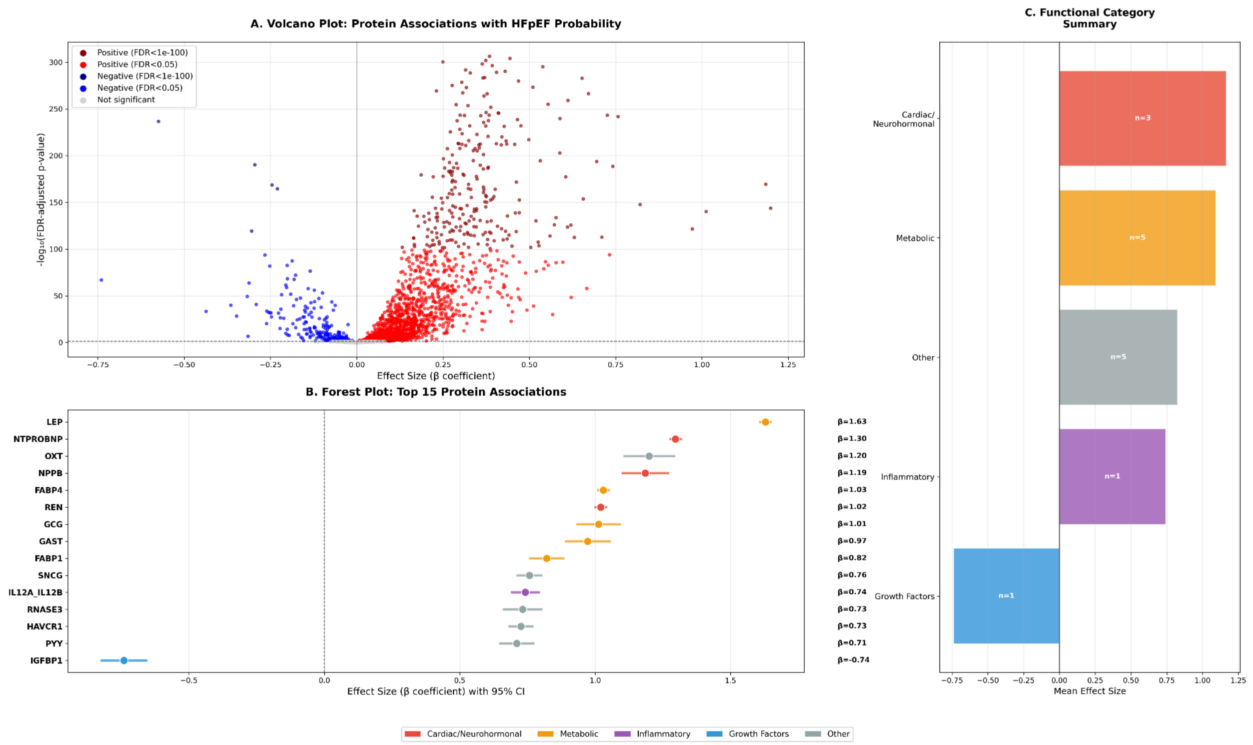


**Supplementary Figure 5.** Proteome-wide analysis. **A)** Volcano plot for positive and negative associated proteins with TRIAD-LAB derived HFpEF predicted probabilities. **B)** 15 proteins with the highest association with TRIAD-LAB derived HFpEF predicted probabilities. **C)** Functional categories of associated proteins.

TRIAD-ECG

There were 2923 proteins available for analysis in the UKB for TRIAD-ECG UKB participants. Our TRIAD-ECG proteome-wide association study revealed significant associations between HFpEF and 1113 plasma proteins (adjusted $p<0.05$).

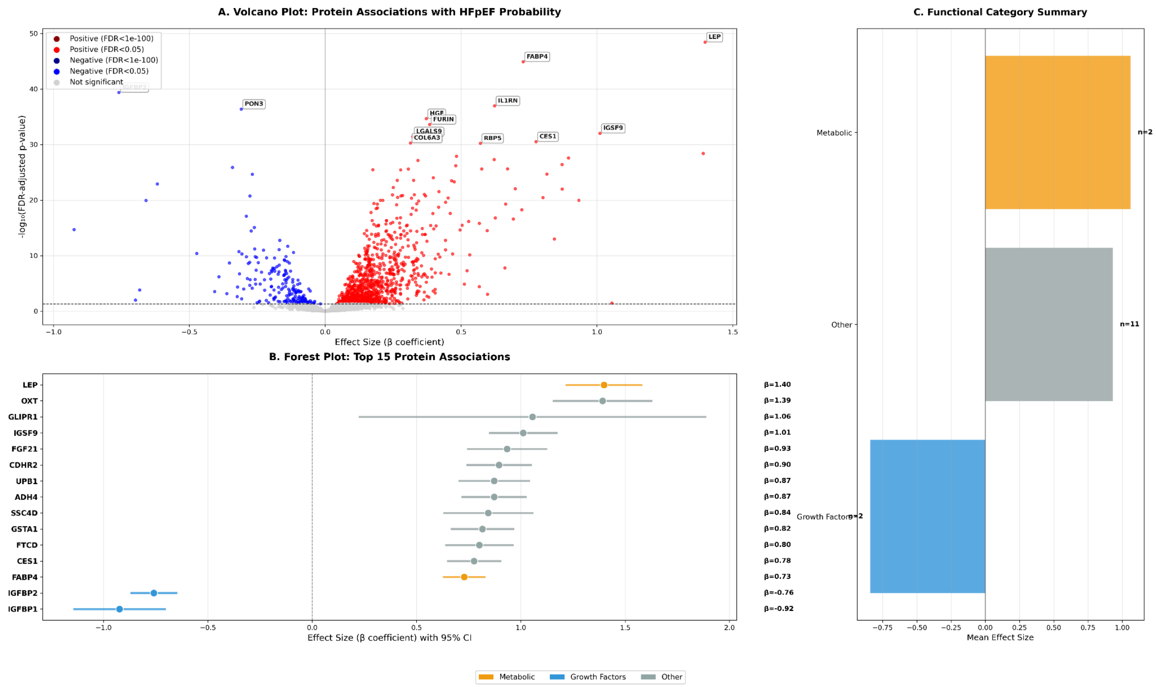


**Supplementary Figure 6** Proteome-wide analysis from TRIAD-ECG

TRIAD-CMR

There were 2923 proteins available for analysis in the UKB for TRIAD-CMR UKB participants. Our TRIAD-CMR proteome-wide association study revealed significant associations between HFpEF and 1456 plasma proteins (adjusted p<0.05).

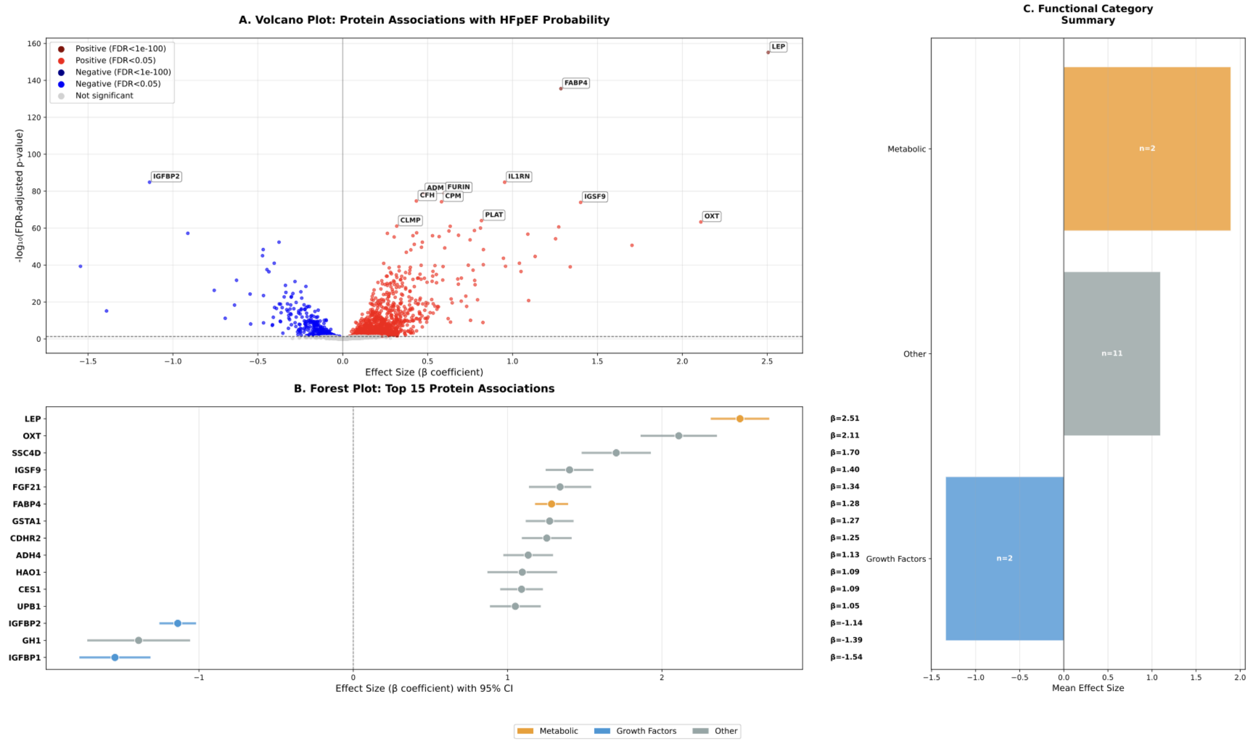

**Supplementary Figure 7** Proteome-wide analysis from TRIAD-CMR

*Colocalization*

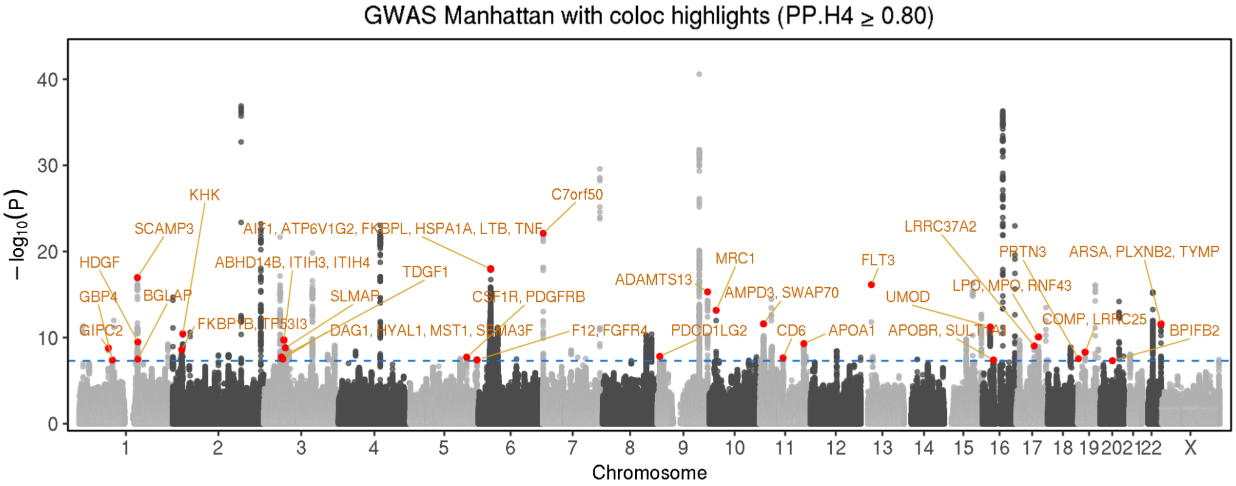


**Supplementary Figure 8.** GWAS Manhattan colocalization plot displaying where predicted probability (PP) is > 0.80 for TRIAD-LAB.

*Protein Quantitative Trait Loci Mendelian Randomization*

TRIAD-LAB

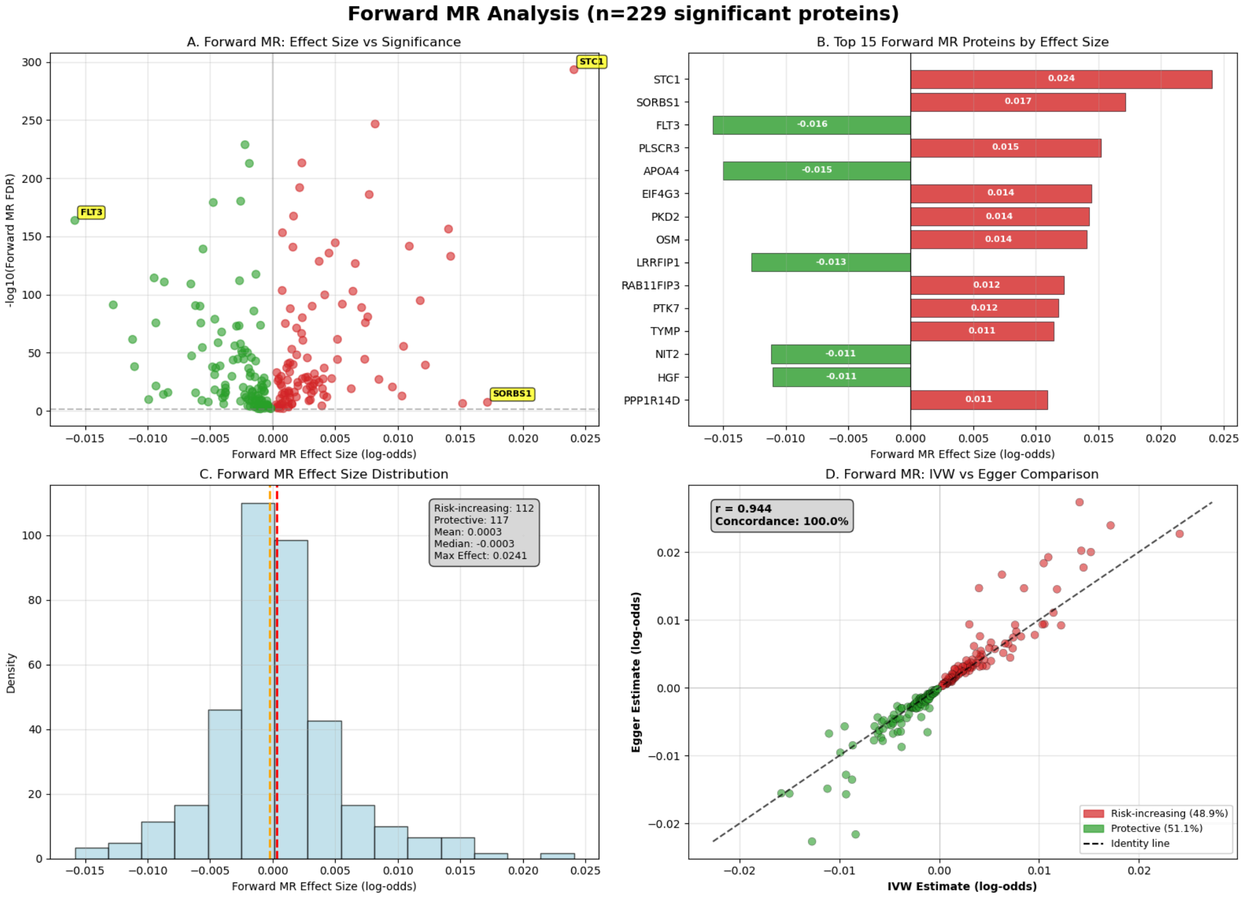


**Supplementary Figure 9.** Forward Mendelian Randomization analysis results for TRIAD-LAB.

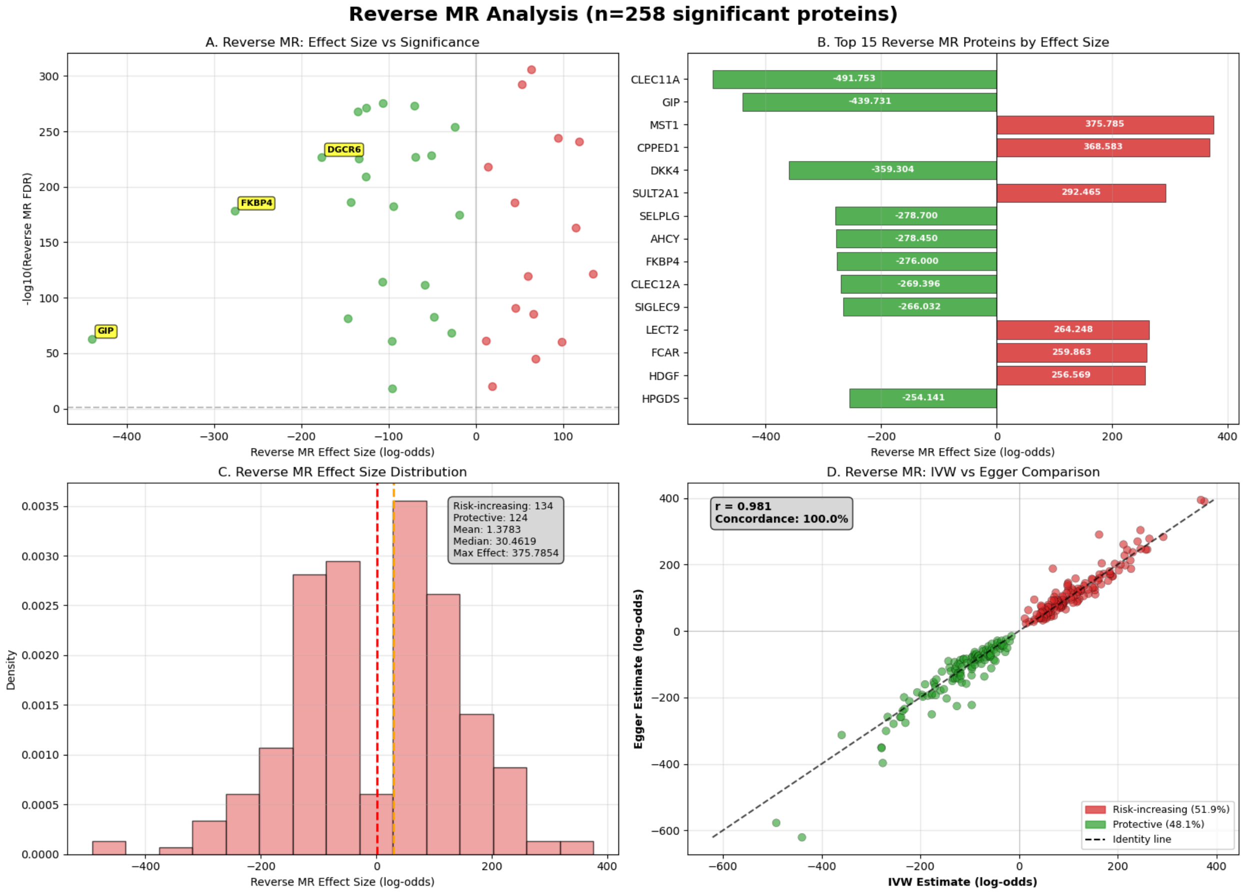


**Supplementary Figure 10.** Reverse Mendelian Randomization analysis results for TRIAD-LAB.

TRIAD-ECG

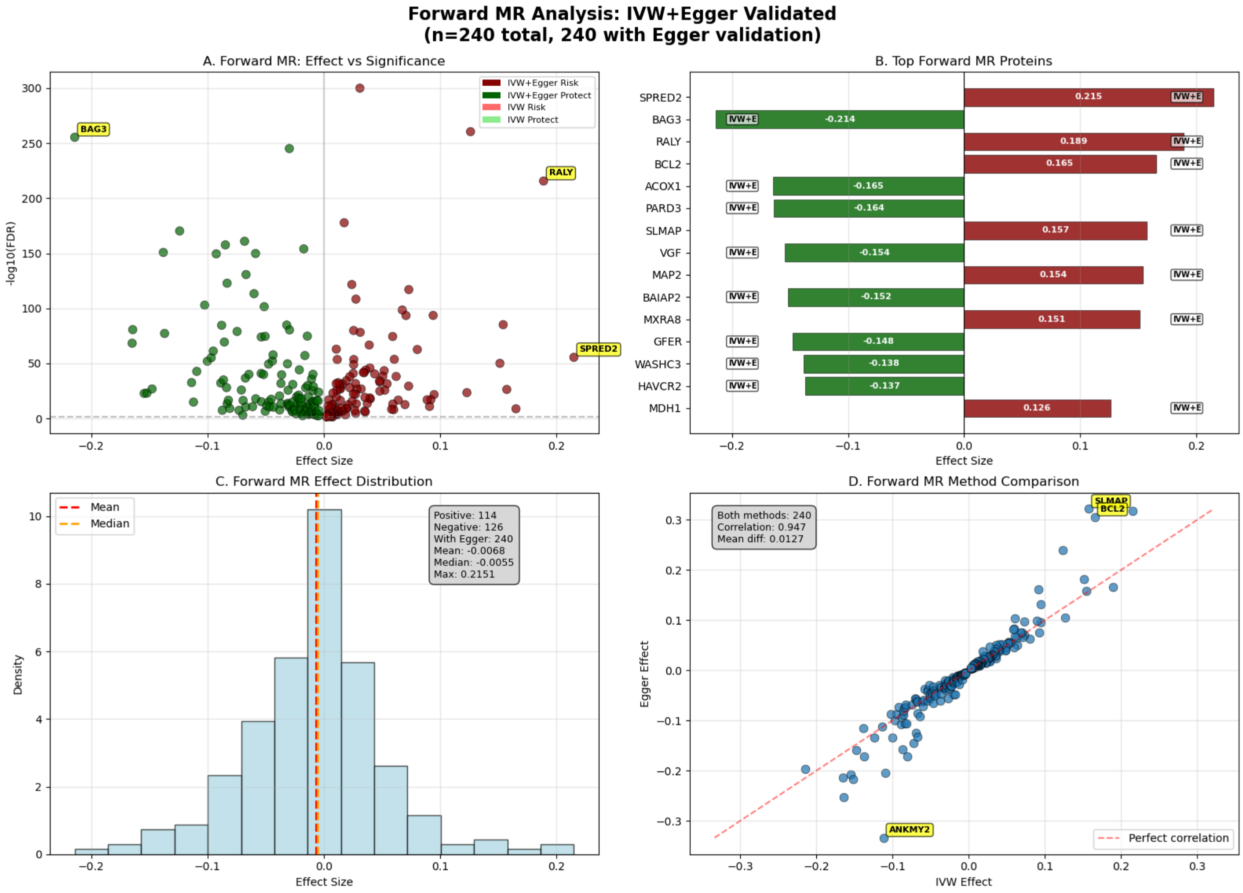


**Supplementary Figure 11** Forward pQTL MR for TRIAD-ECG

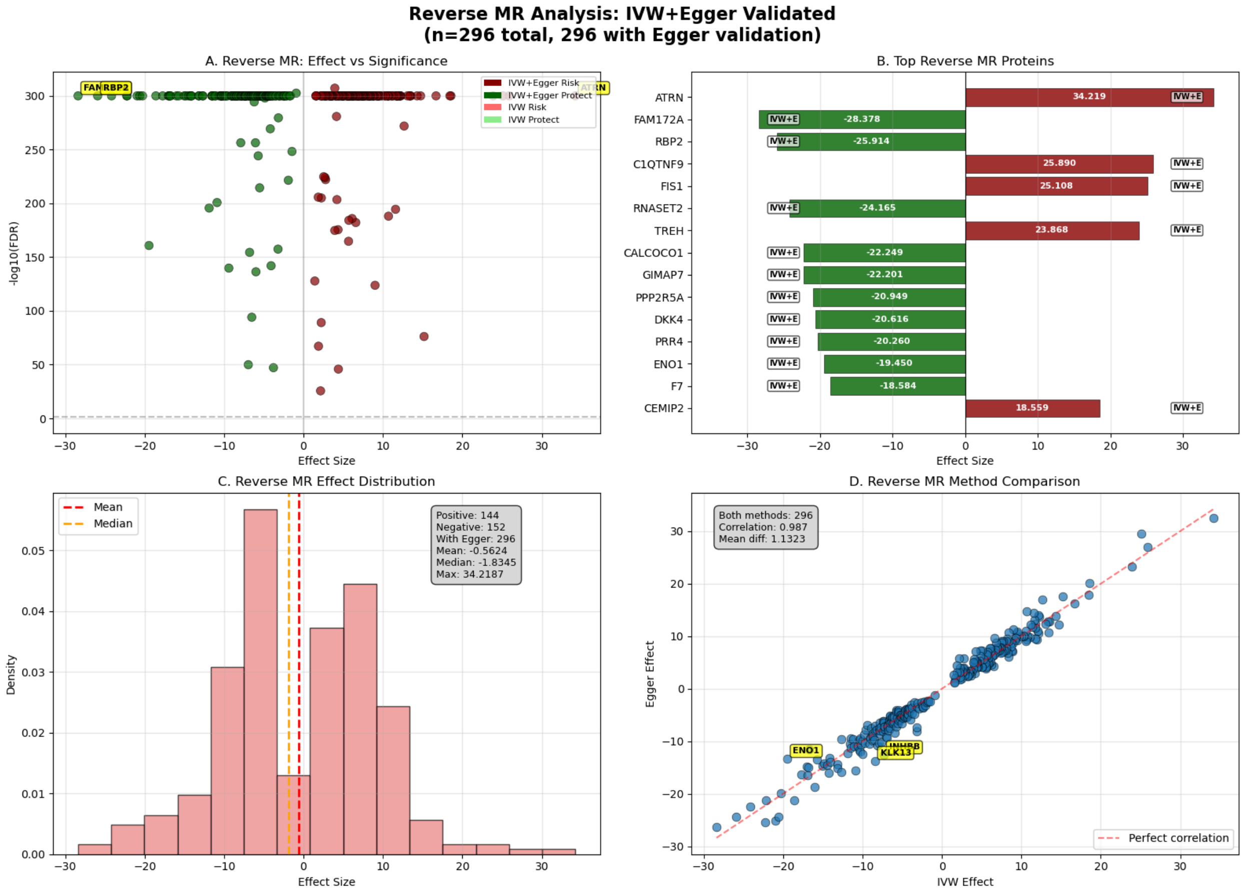


**Supplementary Figure 12** Reverse pQTL MR for TRIAD-ECG

TRIAD-CMR

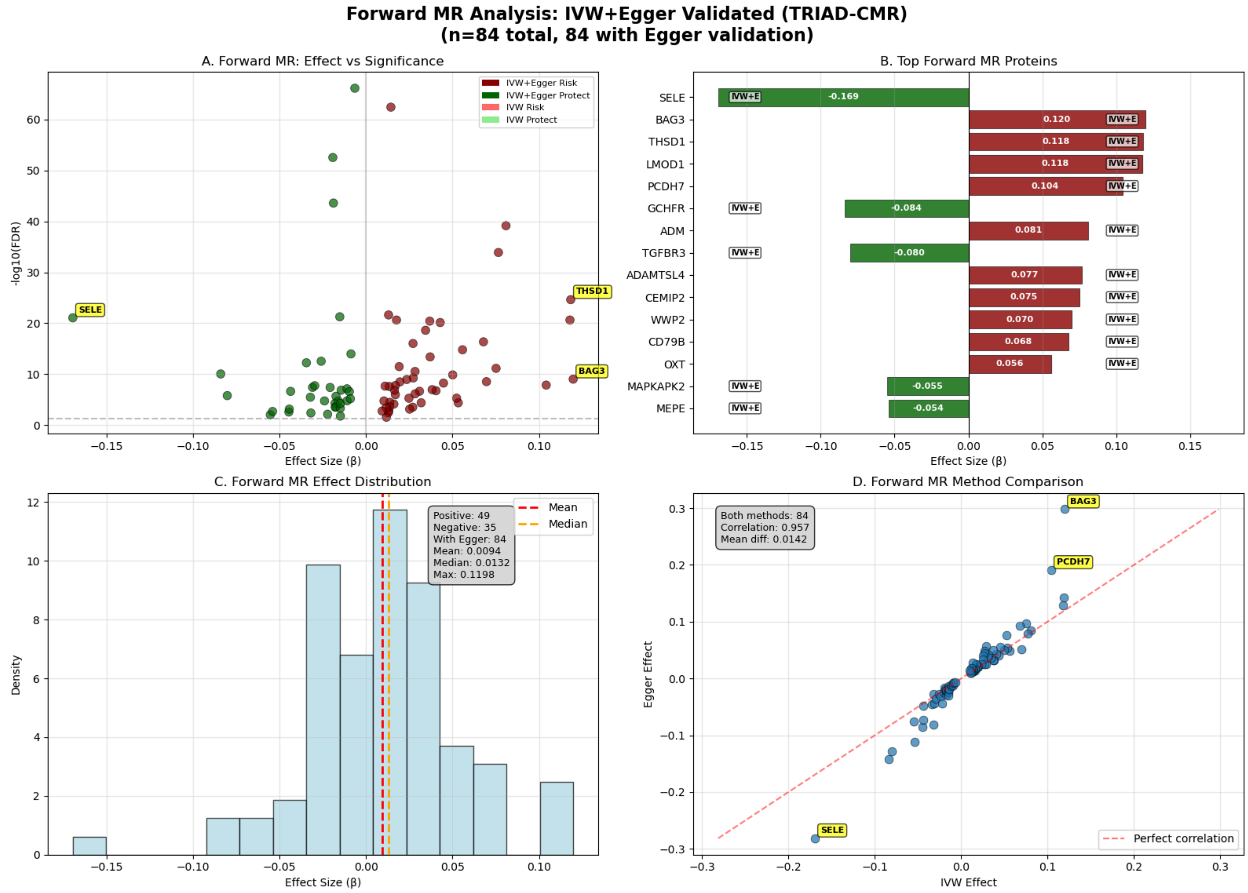


**Supplementary Figure 13** Forward pQTL MR for TRIAD-CMR

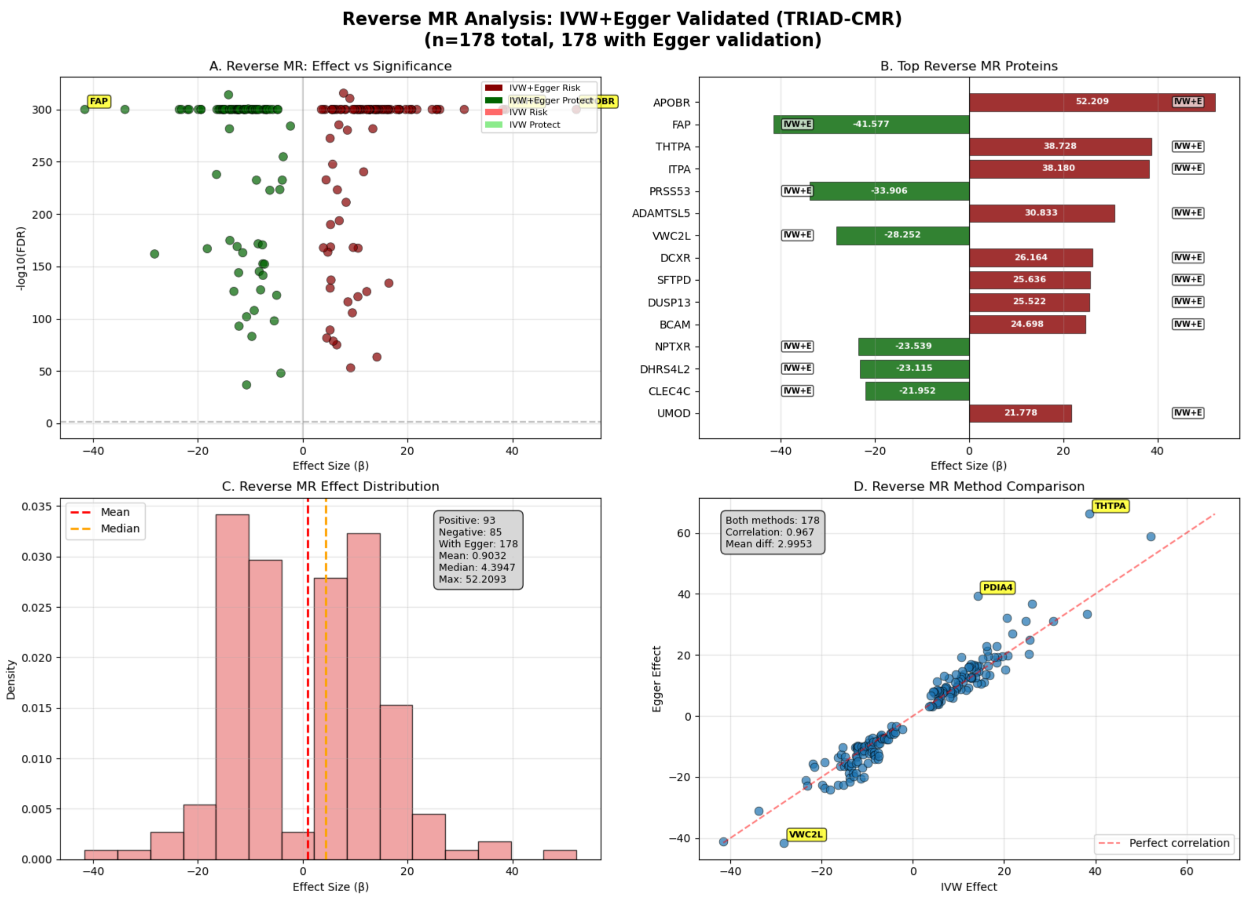


**Supplementary Figure 14** Reverse pQTL MR for TRIAD-CMR

*Systematic prioritization of gene–protein pathways.*

TRIAD-LAB

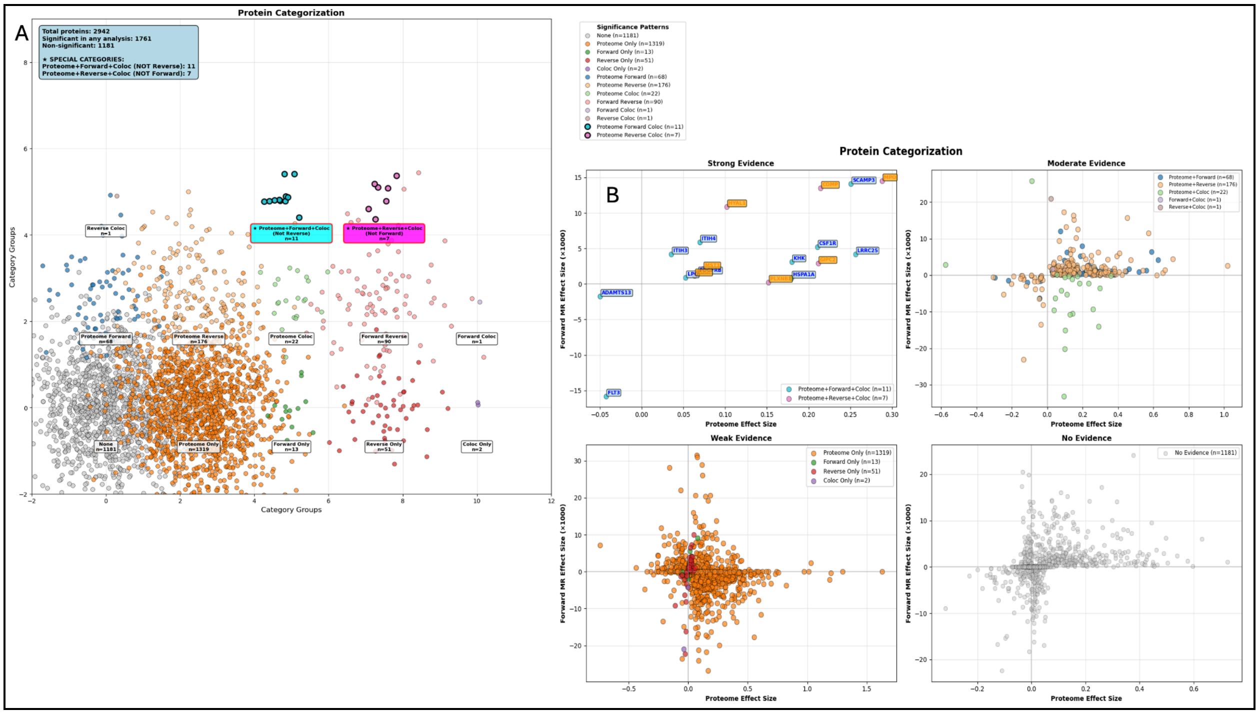


**Supplementary Figure 15** TRIAD-LAB: Systematic priorization of gene-protein pathways. Levels of Evidence across proteome-wide analysis, pQTL MR (IVW, and MR Egger), and co-localization.

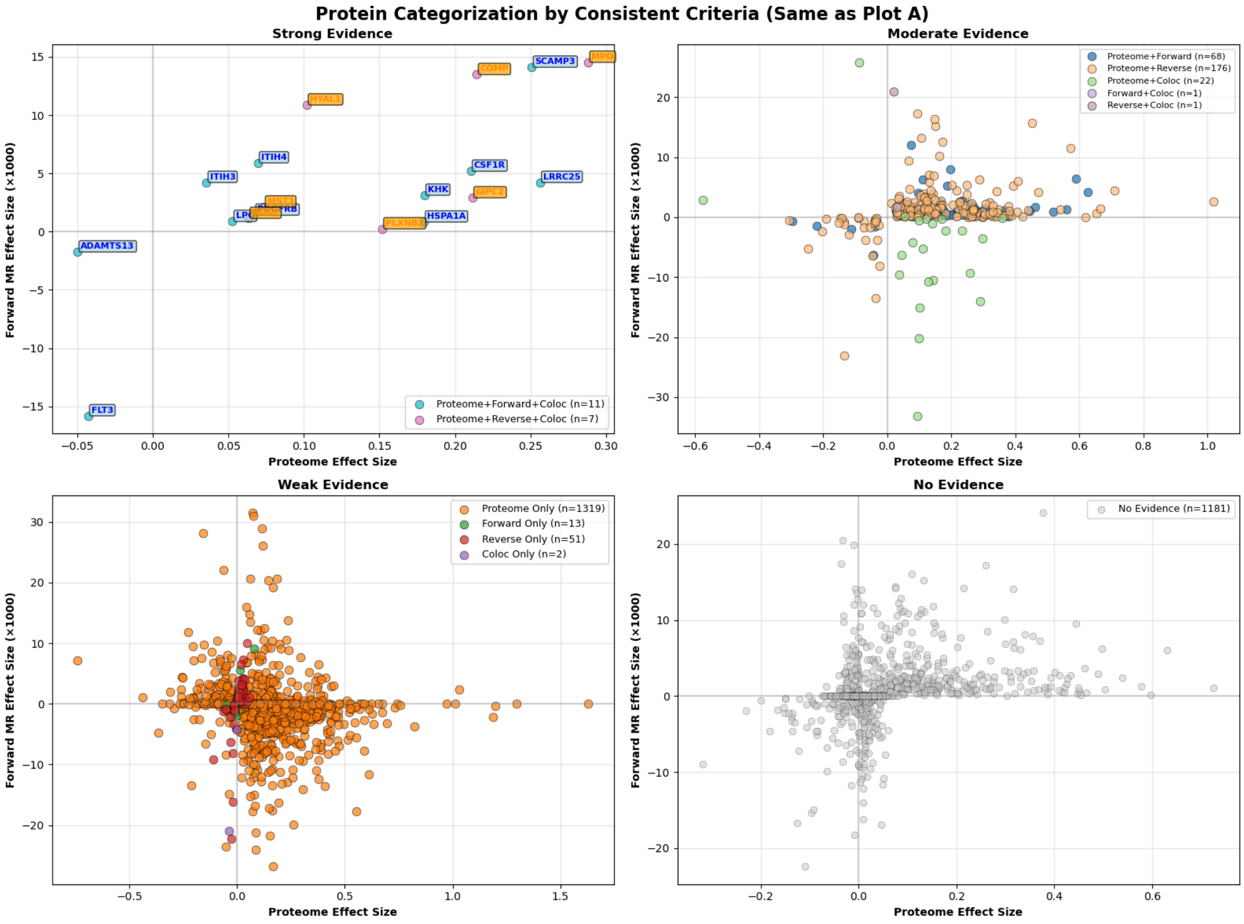


**Supplementary Figure 16** TRIAD-LAB: Enlarged figure of Supplementary Figure 10-B. Systematic priorization of gene-protein pathways. Levels of Evidence across proteome-wide analysis, pQTL MR (IVW, and MR Egger), and co-localization.

TRIAD-ECG

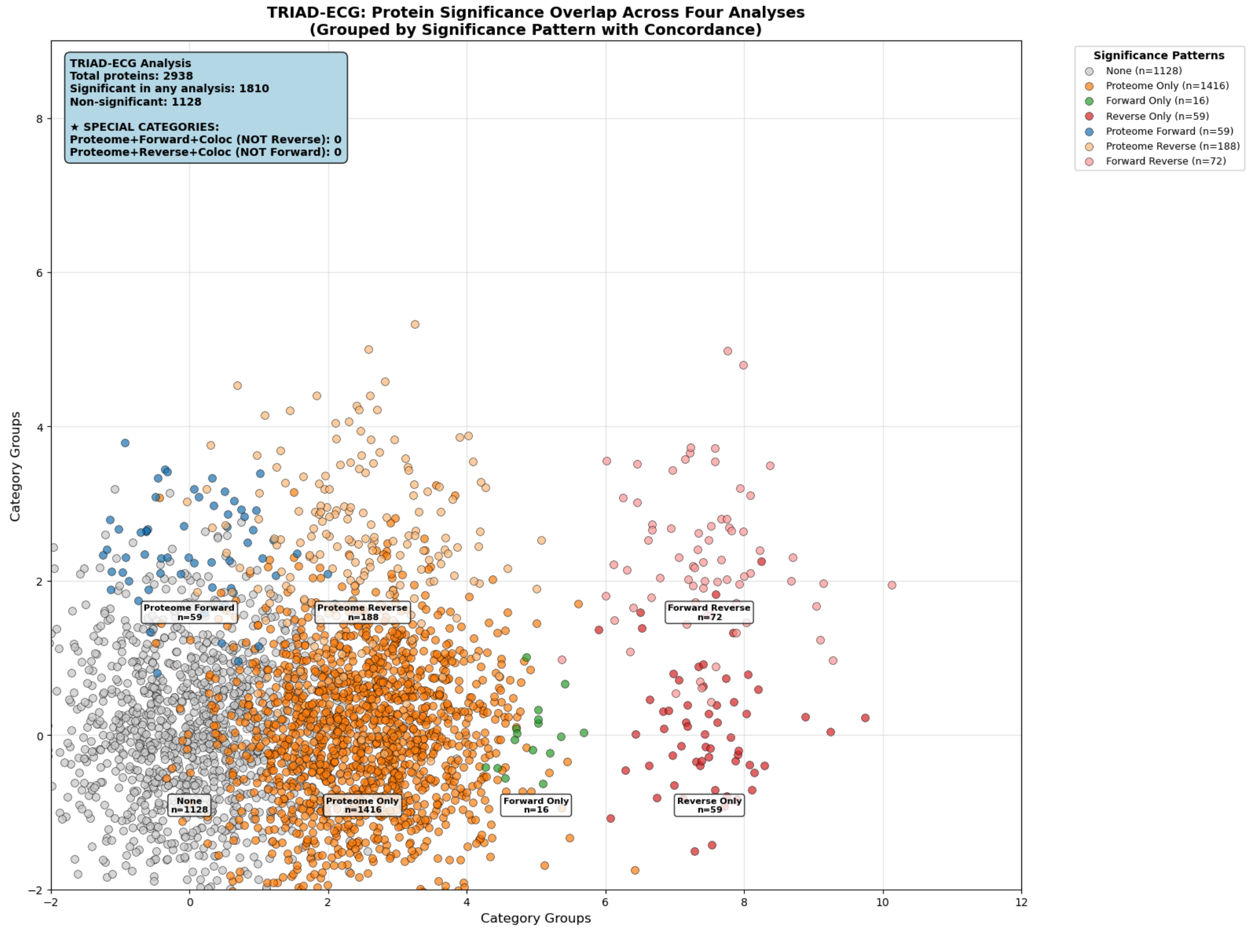


**Supplementary Figure 17** TRIAD-ECG: Systematic priorization of gene-protein pathways. Levels of Evidence across proteome-wide analysis, pQTL MR (IVW, and MR Egger), and co-localization.

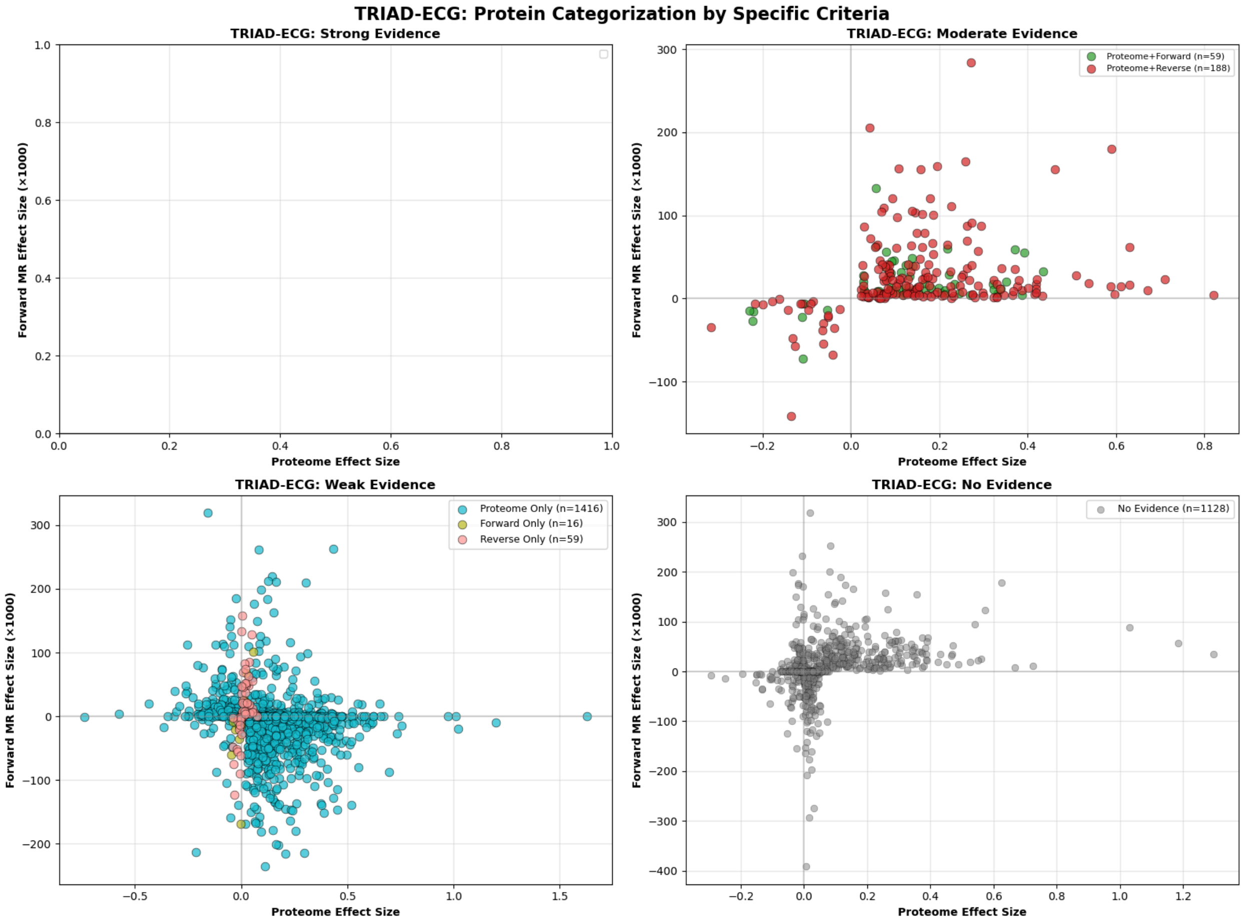


**Supplementary Figure 18** TRIAD-ECG: Systematic priorization of gene-protein pathways. Levels of Evidence across proteome-wide analysis, pQTL MR (IVW, and MR Egger), and co-localization.

TRIAD-CMR

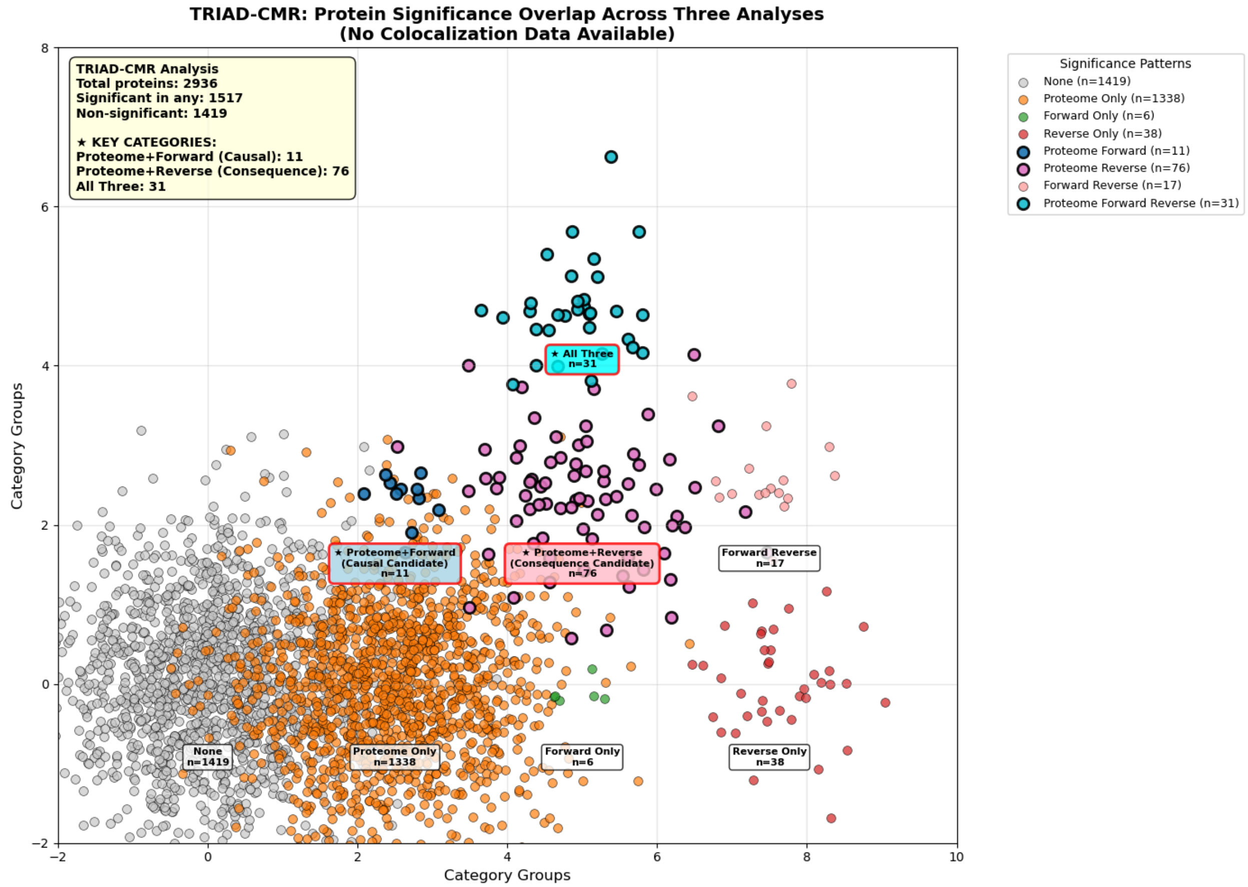


**Supplementary Figure 19** TRIAD-CMR: Systematic priorization of gene-protein pathways. Levels of Evidence across proteome-wide analysis, pQTL MR (IVW, and MR Egger), and co-localization.

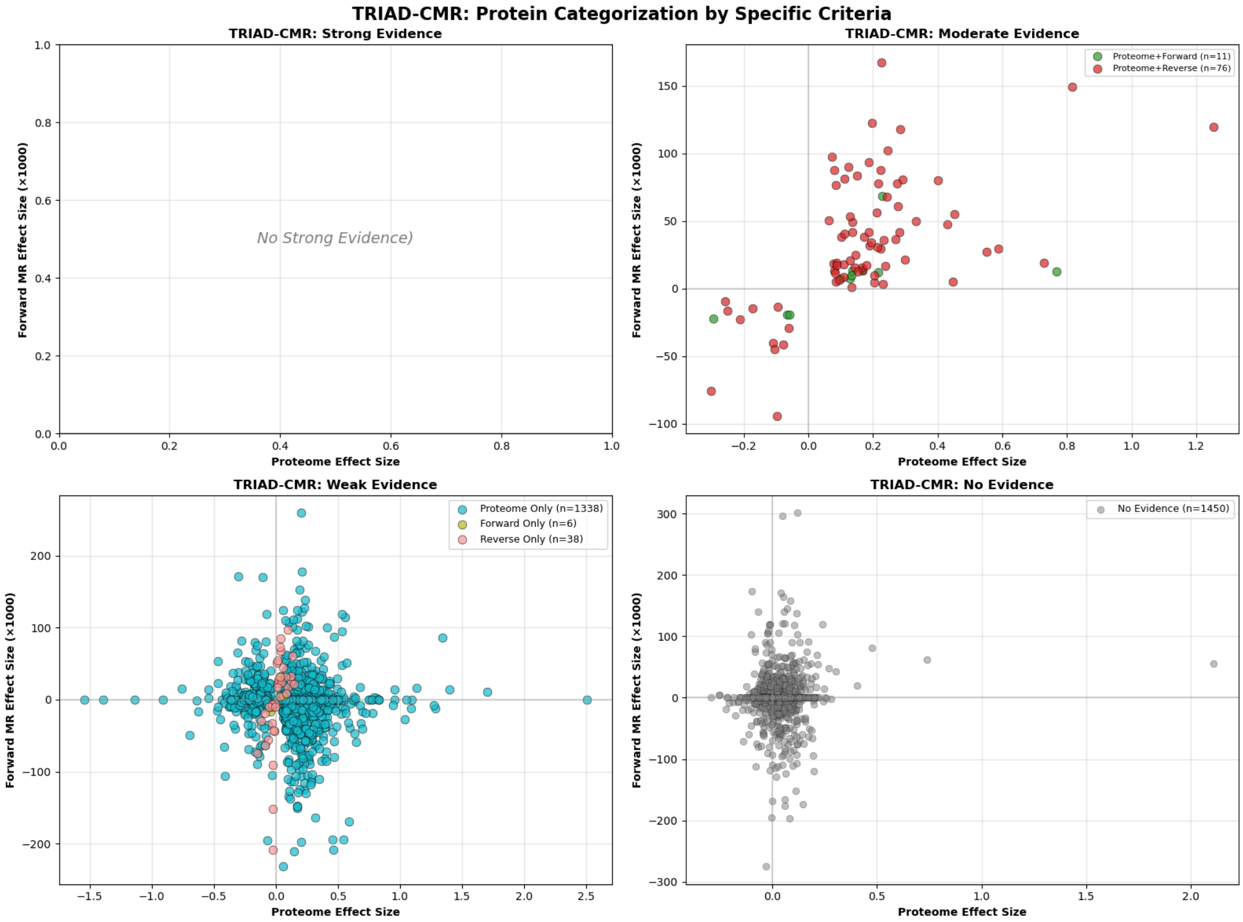


**Supplementary Figure 20** TRIAD-CMR: Systematic priorization of gene-protein pathways. Levels of Evidence across proteome-wide analysis, pQTL MR (IVW, and MR Egger), and co-localization.

*Clinical Validation of FLT3*

| Parameter | Pre | Post | Δ | Adj Δ | % change from baseline | P-value (adjusted) |
|---|---|---|---|---|---|---|
| LVEF (%) | 62.4 | 58.9 | -3.5 | -2.58 | 6 | 0.104 |
| Lat e' (cm/sec) | 11.7 | 8.7 | -3 | -2.54 | 26 | <0.001* |
| Medial e' (cm/sec) | 8.5 | 6.4 | -2.1 | -1.75 | 24 | <0.001* |
| Mitral E/A | 1.1 | 1.1 | -0.1 | 0.04 | 6 | 0.668 |
| LAVi (ml/m2) | 29.1 | 30.1 | 1 | 0.99 | 3 | 0.659 |
| E/e' ave | 9.1 | 10 | 0.9 | 1.24 | 10 | 0.036* |
| Tricuspid Velocity (cm/sec) | 238.8 | 239.1 | 0.3 | 6.52 | 0 | 0.682 |

**Extended Table 1**: Change in LVEF, and diastolic markers before and after FLT3 inhibitor use, both unadjusted (Δ) and adjusted (Adj Δ) for age, time and anthracycline use.

*All open source code*

Phenotyping code

- To extract LVEF and diastolic measures: https://github.com/jackosullivanoxford/HFpEF_phenotyping_and_multiomics/code_to_phenotype_HFpEF

TRIAD-HFpEF Multimodal model code and model weights

- *TRIAD-ECG:* https://github.com/jackosullivanoxford/TRIAD_ECG_HFPEF/tree/main
    - *TRIAD-ECG model weights:* https://huggingface.co/jackosullivan/TRIAD_ECG_HFPEF/tree/main
- *TRIAD-CMR:* https://github.com/Google-Health/genomics-research/tree/main/triad-hfpef
- *TRIAD-LAB:* https://github.com/jackosullivanoxford/TRIAD_LAB_HFpEF `
    - *TRIAD-LAB model weights:* https://huggingface.co/jackosullivan/TRIAD_LAB_HFPEF/tree/main

Validation of TRIAD-HFpEF assigned codes

- https://github.com/jackosullivanoxford/HFpEF_phenotyping_and_multiomics/HFpEF_validation_survival
- LVEF and Cox-survival: https://github.com/jackosullivanoxford/HFpEF_phenotyping_and_multiomics/blob/main/Association_with_CMR_tabular_and_LVEF

Proteome-wide association code

- https://github.com/jackosullivanoxford/HFpEF_phenotyping_and_multiomics/Proteome_wide_analysis

pQTL MR code:

- github.com/jackosullivanoxford/HFpEF_phenotyping/pQTL_HFpEF_MR_IVW method
- github.com/jackosullivanoxford/HFpEF_phenotyping/pQTL_HFpEF_MR_Egger_method.

Colocalization analysis

- https://github.com/jackosullivanoxford/HFpEF_phenotyping_and_multiomics/colocalization

*Supplementary tables*

*All open source GWAS Summary Statistics*

- GWAS Summary statistics TRIAD-ECG: https://zenodo.org/records/17945999
- GWAS Summary statistics TRIAD-CMR: https://zenodo.org/records/17946070
- GWAS Summary statistics TRIAD-LAB: https://zenodo.org/records/17946052
- Google Cloud Storage: https://console.cloud.google.com/storage/browser/brain-genomics-public/research/triad-hfpef
